# The current burden of Japanese encephalitis and the estimated impacts of vaccination: Combining estimates of the spatial distribution and transmission intensity of a zoonotic pathogen

**DOI:** 10.1101/2021.04.08.21255086

**Authors:** Sean M. Moore

**Affiliations:** Department of Biological Sciences and Eck Institute for Global Health, University of Notre Dame, Notre Dame, IN 46556

## Abstract

Japanese encephalitis virus (JEV) is a major cause of neurological disability in Asia and causes thousands of severe encephalitis cases and deaths each year. Although Japanese encephalitis (JE) is a WHO reportable disease, cases and deaths are significantly underreported and the true burden of the disease is not well understood in most endemic countries. Here, we first conducted a spatial analysis of the risk factors associated with JE to identify the areas suitable for sustained JEV transmission and the size of the population living in at-risk areas. We then estimated the force of infection (FOI) for JE-endemic countries from age-specific incidence data. Estimates of the susceptible population size and the current FOI were then used to estimate the JE burden from 2010 to 2019, as well as the impact of vaccination. Overall, 1.15 billion (range: 982.1-1543.1 million) people were estimated to live in areas suitable for endemic JEV transmission, which represents 28.0% (range: 24.0-37.7%) of the over four billion people living in countries with endemic JEV transmission. Based on the baseline number of people at risk of infection, there were an estimated 45,017 (95% CI: 13,579-146,375) JE cases and 16,319 (95% CI: 1,804-60,041) deaths in 2019. Estimated incidence declined from 61,879 (95% CI: 18,377-200,406) cases and 22,448 (95% CI: 2,470-83,588) deaths in 2010, largely due to increases in vaccination coverage which have prevented an estimated 214,493 (95% CI: 75,905-729,009) cases and 78,544 (95% CI: 8,243-325,755) deaths over the past decade. India had the largest estimated JE burden in 2019, followed by Bangladesh and China. From 2010-2019, we estimate that vaccination had the largest absolute impact in China, with 142,471 (95% CI: 56,208-484,294) cases and 52,338 (95% CI: 6,421-185,285) deaths prevented, while Taiwan (91.1%) and Malaysia (80.5%) had the largest percent reductions in JE burden due to vaccination. Our estimates of the size of at-risk populations and current JE incidence highlight countries where increasing vaccination coverage could have the largest impact on reducing their JE burden.

**Author Summary:** Japanese encephalitis is a vector-transmitted, zoonotic disease that is endemic throughout a large portion of Asia. Vaccination has significantly reduced the JE burden in several formerly high-burden countries, but vaccination coverage remains limited in several other countries with high JE burdens. A better understanding of both the spatial distribution and the magnitude of the burden in endemic countries is critical for future disease prevention efforts. To estimate the number of people living in areas within Asia suitable for JEV transmission we conducted a spatial analysis of the risk factors associated with JE. We estimate that over one billion people live in areas suitable for local JEV transmission. We then combined these population-at-risk estimates with estimates of the force of infection (FOI) to model the national-level burden of JE (annual cases and deaths) over the past decade. Increases in vaccination coverage have reduced JE incidence from over 60,000 cases in 2010 to 45,000 cases in 2019. We estimate that vaccination has prevented over 214,000 cases and 78,000 deaths in the past decade. Our results also call attention to the countries, and high-risk areas within countries, where increases in vaccination coverage are most needed.

## Introduction

Japanese encephalitis virus (JEV) is a mosquito-transmitted flavivirus that is endemic in a large portion of South and Southeast Asia, as well as parts of the Western Pacific. Previous estimates have suggested that over three billion people live in countries with JEV transmission, resulting in an estimated 68,000 clinical cases and over 13,000 deaths annually [1]. Fewer than 1% of JEV infections in humans are symptomatic, but the case-fatality rate among clinical encephalitis cases is 15-30% and up to 50% of surviving encephalitis cases experience long-term neurological or psychiatric sequelae [2]. Japanese encephalitis is considered a zoonotic disease because humans are dead-end hosts for the pathogen [3]. Therefore, a zoonotic cycle of transmission between JEV-competent mosquitos and either a wildlife or domestic animal reservoir is required for persistence (or frequent reintroduction) in JE-endemic areas [3].

Several JEV vaccines have been in use since the 1930s, with the prequalified live attenuated SA14-14-2 vaccine used most frequently in endemic countries [4]. Several countries that formerly had a high burden of JE, such as Japan, Taiwan, and South Korea, have reduced that burden to almost zero mainly through high vaccination coverage [2]. These successes suggest that although vaccination does not remove the zoonotic source of JEV transmission, spillover to humans can be significantly reduced via large-scale vaccination. Increased funding for JEV vaccination in recent years, including support from Gavi, the Vaccine Alliance (Gavi) beginning in 2013, provides an opportunity to extend the successes experienced in higher-income countries to other countries that still experience a significant burden. However, with over three billion people living in JEV-endemic countries, existing resources need to be targeted towards the most at-risk populations. A systematic review estimated that 68,000 JE cases and 13,000-20,000 deaths occur annually in countries where JEV circulates [1]. However, due to a lack of detailed surveillance and reporting, this burden estimate was obtained by extrapolating incidence rates from twelve different epidemiological studies to the entire JE-endemic region. Because local transmission to humans requires environmental conditions capable of sustaining an enzootic cycle, or at least sporadic zoonotic outbreaks that can spillover to humans, the risk of infection can vary substantially within each endemic country.

Identifying where spillover transmission is likely to occur is an important component to accurately estimating the burden of disease and the impact of vaccination for zoonotic diseases such as JE. For a vector-transmitted, zoonotic pathogen such as JEV, spatial heterogeneity in transmission intensity within a country can be quite high since the chances of spillover will be zero where either competent animal hosts or vectors are absent (although sporadic spillovers may occur in areas where migratory waterfowl serve as occasional hosts or certain zoophilic vectors occasionally bite humans). Therefore, even in regions with a high mean force of infection (FOI), a portion of the population will likely be at a zero- or low-risk of infection. JEV transmission is typically highest in rural areas, and is associated with irrigated rice paddies [5]. The presence of competent reservoir and amplifying hosts is also important for spillover intensity [3]. The increased availability of high resolution spatial datasets cataloging not only environmental and ecological conditions, but also land use and animal distributions, can be used to identify areas suitable for sustained JEV transmission, as well as the size of the potentially susceptible human population.

While several migratory bird species have been implicated as reservoir hosts [3], the majority of transmission to humans is believed to be in areas where domestic livestock serve as reservoirs or amplifying hosts [5]. Cattle and horses are believed to be dead end hosts for the virus, while domestic pigs are good zoonotic hosts because they can infect mosquitoes [3]. In addition, the high turnover in domestic pig populations introduces new susceptible individuals frequently and can seasonally amplify transmission. Domestic fowl (chickens and particularly ducks) have also been identified as possible sources of transmission in some areas [6]. Cases are often concentrated in areas where humans and their livestock live in close proximity to natural wetlands or irrigated farmland, including rice paddies [3].

In addition to determining the proportion of the population that is currently susceptible and at risk of infection, estimating the disease burden requires an estimate of the transmission intensity in endemic areas where the population is at risk of infection. Reported disease incidence may not be an accurate measure of transmission intensity because higher transmission rates can lower the average age of infection rather than increasing incidence rates [7]. Instead, transmission intensity can be measured as the force of infection (FOI) experienced per susceptible individual in the population, which can be estimated from age-specific incidence in endemic settings [8]. Here we provide an example of how estimates of the FOI for JE-endemic countries can be used to estimate the current transmission intensity at a national or sub-national level. Combining these estimates from epidemiological data with a spatial analysis of the population at risk of infection allows us to model current JE incidence and estimate the impact of vaccination on disease incidence over the past decade as vaccination coverage has increased in several countries.

## Methods

### Estimating size of at-risk population

To estimate the number of people living in areas with likely JEV transmission we used several different spatial datasets representing factors associated with the maintenance of the zoonotic JEV cycle and subsequent spillover to humans. To identify areas with suitable habitat for JEV transmission we used a fine-scale map of the extent of lowland rice production in Asia modeled from satellite data by the International Rice Research Institute (IRRI) [9] (SI Fig 1). This dataset was combined with a new map of seasonal and permanent wetlands in the tropics and subtropics at a 231×231 m spatial resolution generated using a hydrological model and satellite-derived estimates of soil moisture to represent water flow and surface wetness [10] (SI Fig 1). A combined map of the areas classified as either wetland or under rice cultivation was used as a baseline for suitable JEV transmission. The number of people living in these areas was calculated using maps of human population density at a 1 km^2^ resolution for 2020 from Worldpop [11].

To further refine our estimates, we incorporated the modeled spatial distribution of the main JEV mosquito vector, *Culex tritaeniorhynchus*, based on environmental suitability at a 5×5 km resolution [12]. Population-at-risk estimates were calculated for all rice and wetland areas where the probability of occurrence for *C. tritaeniorhynchus* was ≥ 0.25. Although *C. tritaeniorhynchus* is considered the main vector for JEV, the virus has been isolated from over thirty other mosquito species [5]. In addition, epidemiological data from several locations, such as Bali, Indonesia, suggest that JEV transmission intensity and spillover to humans can be high even where environmental suitability for *C. tritaeniorhynchus* is predicted to be low. Areas of low environmental suitability for *C. tritaeniorhynchus* with known circulation of JEV appear to be concentrated in the hotter, tropical regions of countries such as Malaysia and Indonesia. An earlier attempt at modeling the spatial distribution of *C. tritaeniorhynchus* did predict higher probabilities of occurrence in hotter, wetter tropical areas [13] compared to the updated distribution model of Longbottom et al. [12]. In addition, a recent analysis of several arboviruses transmitted by *Culex* species, including two flaviviruses, found that the optimal temperatures for viral transmission were between 23 *−*26^*°*^C [14]. Therefore, to capture areas where the environmental suitability for *C. tritaeniorhynchus* may be underestimated, or where conditions are ideal for transmission by other mosquito species, we also included areas where the annual mean minimum temperature was ≥20^*°*^C and annual rainfall exceeded 150 cm, because these thresholds captured the known locations of JEV occurrence in Indonesia, Malaysia, and the Philippines where the predicted environmental suitability for *C. tritaeniorhynchus* is low (SI Fig 2).

Due to the strong association between JEV transmission to humans and the presence of domestic pigs, it is likely that areas with high domestic pig densities are capable of experiencing high transmission intensities. However, relatively high JE incidence rates have been observed in areas of Bangladesh [15] and Indonesia [16] that have few pigs because the local population is majority Muslim. In addition to pigs, domestic fowl (particularly ducks) have been associated with JEV transmission [5] and they can be present in high densities in the regions of Bangladesh, and on the Indonesian islands of Sumatra and Java, where JE cases have been reported despite the lack of local domestic pig populations. To best represent a more conservative estimate of the size of the at-risk population, we calculated the number of people living in areas with the appropriate habitat type (rice and wetland), and suitable environmental conditions for the mosquito vector, where the combined density of pigs and ducks exceeded a certain threshold. Estimated cattle, pig, chicken, and duck population densities at a 1×1 km resolution were taken from the Global Distribution of Livestock dataset [17]. The initial threshold for JE suitability was a combined local density of 2 pigs or ducks per square km, with sensitivity to this threshold value tested by ranging the threshold from 1-10 per sq km (SI Fig 3). The size of the population at risk of infection was identified as those living in areas with a combined pig and duck population above the minimum threshold, a land use type of wetland or rice production, and environmental conditions suitable for the mosquito vector. In addition, as part of our sensitivity analysis we used an upper estimate for the size of the population at risk based on only the habitat type (wetland or rice production).

### Epidemiological and vaccination data

To estimate the FOI in each JE-endemic country at the national level, we conducted a non-systematic review of publications that reported age-specific JE incidence data. The search was restricted to studies reporting either confirmed or probable JE cases, excluding studies that only reported acute encephalitis cases (AES) because the fraction of AES attributable to JE varies considerably both spatially and temporally [1]. Details of the studies included in our analysis are provided in SI Table 1. For countries where at least one study was available prior to the introduction of vaccination, any studies that were conducted after the introduction of vaccination were excluded to avoid having to estimate both FOI and vaccination coverage simultaneously. As a result, we excluded incidence data from 2007 in Nepal [18] and after 2001 from Sarawak, Malaysia [19].

For several countries (China, India, Japan, Malaysia, Sri Lanka, South Korea, Taiwan, Thailand, and Vietnam), incidence data was only available for time periods after the introduction of JEV vaccine into at least part of the study area, so for these countries we simultaneously estimate both FOI and vaccination coverage. Initial estimates of vaccination coverage in these countries were obtained from literature sources identified by Quan et al. [20] and routine vaccination coverage estimates provided by WHO-UNICEF joint reporting [21]. A list of references used to generate preliminary vaccination estimates for each study are provided in SI Table 1. Population age distributions for each study were assumed to match the national age distributions in the UN World Population Prospects demographics dataset [22].

### Estimating the force of infection

In an unvaccinated population, the FOI can be estimated from either serological or case data using a catalytic model [7]. For a constant FOI (*λ*), the proportion of the population that remains susceptible at age *a* is calculated using the catalytic model equation:

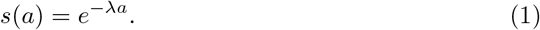

Due to the lack of longitudinal studies on JE transmission dynamics, we assumed that *λ* does not vary with age. While there are likely to be age- and sex-specific risks of exposure to JEV, age differences in FOI rates are likely to be smaller than those observed in many vaccine-preventable childhood diseases where assortative mixing among school-age children leads to significant differences in age-specific FOIs. A constant FOI can be directly estimated from seroprevalence data by assuming that *s*(*a*) is equivalent to the fraction of seronegative individuals at age *a*.

To estimate the FOI from age-structured incidence data we must estimate both the probability that an individual remains uninfected until age *a* and the probability that an uninfected individual of age *a* will become infected at that age. For a population that is stratified into *M* age groups, with age group *j* spanning in age from 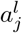 to 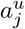, the proportion of the population that will become infected in age group *j*, Γ_*j*_, is the integral of the probability of being susceptible at age *a* multiplied by the FOI:

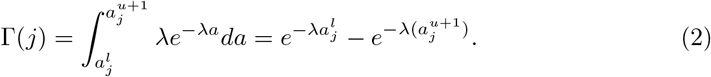

If a portion of the population has been vaccinated, then the probability that an individual remains susceptible to infection at age *a* from equation (1) is modified to:

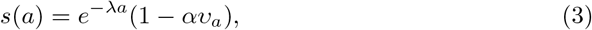

where *α* is the vaccine efficacy and *υ*_*a*_ is the probability of being vaccinated by age *a*. This equation assumes that vaccine-derived immunity does not wane over time. The proportion of individuals infected at age *a* based on *λ* and the proportion who have not been previously infected or vaccinated is represented as:

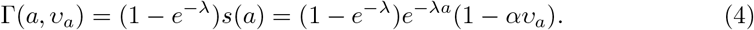

We assume that the proportion of individuals who are infected in age group *j* spanning in age from 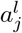 to 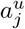 can then be calculated as

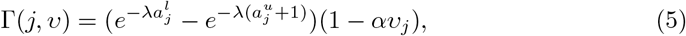

where *υ*_*j*_ is the vaccination coverage for age group *j*.

The FOI was estimated for each study using an MCMC approach. The observed vector of JE cases per age class in year *t, I*_*t*_, is a vector of length equal to the number of reported age classes, where each element, *I*_*j,t*_ is the number of cases observed in age class *j. I*_*t*_ was modeled as

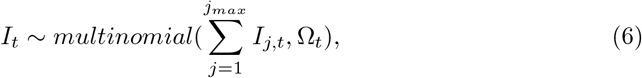

where Ω_*t*_ is a vector of probabilities that depends on the proportion of age class *j* that is infected in year *t*, Γ_*j,t*_, and the proportion of the study population that is in age class *j* in year *t, θ*_*j,t*_,

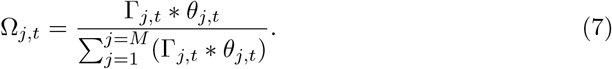

For studies conducted in populations that have been partially vaccinated, the proportion of age group *j* that is infected, Γ_*j,t*_, depends on both *λ* and vaccination coverage rates for age class *j* in year *t, υ*_*j,t*_. In addition to estimating *λ*, we also estimate the vaccination coverage for each age class. Initial values for *υ*_*j,t*_ were obtained from the data sources listed in SI Table 1. We assumed that individuals of age *a* in year *t* were vaccinated as infants with a probability equal to the reported routine vaccination coverage level 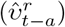 in year *t − a*. Vaccination campaigns occurring between year *t – a* and year *t* were included if an individual was in the target population so that

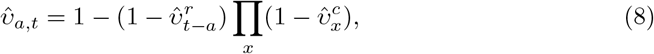

where 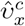 is the reported coverage level in the target population for campaign *x*.

Study-specific force of infection (*λ*) values were estimated by fitting equations (2,6,7) in the absence of vaccination and equations (4,5,6,7) in the presence of vaccination. We used a non-negative, truncated *Normal*(0, 10) prior for *λ*. To incorporate sufficient uncertainty in the priors for the vaccination coverage parameters, *υ*_*a,t*_, we used an informative prior 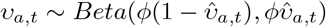 following the approach detailed by Quan et al. [20]. The value 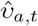 was the initial estimate of the vaccination coverage in age class *a* derived from reported vaccination coverage levels at the national level (see SI Table 1 for data sources). The uncertainty in the vaccination information is represented by setting *ϕ* = 5. Model fitting was done using a Bayesian framework via a Markov chain Monte Carlo (MCMC) approach implemented in STAN using the ‘rstan’ version 2.18.2 package in R [23]. The models were run with four chains of 10,000 iterations each and a 50% burn-in period. Smaller step sizes for the sampling algorithm were set by increasing the adapt delta parameter from the default of 0.8 to 0.99. In addition, the maximum tree depth was increased from 10 to 15. Convergence was assessed using the Gelman-Rubin convergence diagnostic, *R*_*c*_ [24]. Posterior predictive checks were performed by comparing the empirical data to data simulated from the posterior parameter distributions for each dataset. Posterior predictive data was simulated at each iteration, *k*, of the MCMC, with 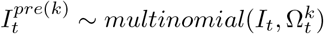. At each iteration, the total observed number of cases, *I*_*t*_, was compared to the total predicted number of cases,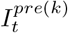. This test statistic was used to calculate a Bayesian p-value 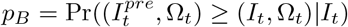, which indicates whether the distribution of the model-generated data was more extreme than the observed data [24].

### Factors associated with the force of infection

Following the estimation of country-specific FOIs from age-specific incidence data, we conducted a regression analysis to determine whether any study-specific or country-specific variables were associated with FOI. The study-specific variables included in the analysis were the start and end years of the study, and whether the study was conducted in an area with a history of JE vaccination. Country-specific variables included the estimated 2020 population size [11], the regional classification and incidence level (very low, low, medium, medium-high, high) from Campbell et al. [1], national per capita gross domestic product (GDP) [25], total land area, the proportion of land area classified as urban [26], the proportion of land area under rice cultivation [9], the mean environmental suitability and the total amount of suitable habitat for *C. tritaeniorhynchus* [12], and the national population sizes and mean population densities of domesticated cows, pigs, ducks, and chickens [17]. In addition, we also included our baseline and low-end estimates of the percentage of the population living in at-risk areas as potential explanatory variables. We examined the Pearson correlation coefficient between the median FOI estimate and each covariate to identify potential explanatory variables. We then performed model selection via a best subsets regression approach using the ‘leaps’ package in R [27]. The total land area was highly correlated with population size, the population sizes of domestic cows, pigs, ducks, and chickens, the amount of area under rice cultivation, and the total amount of habitat for *C. tritaeniorhynchus*. Therefore, of these variables only total land area was retained as a potential explanatory variable during model selection. We conducted an exhaustive search of all possible models with up to ten explanatory variables and used BIC values to select the best model.

### Estimating annual JE burden and vaccine impacts

The annual number of JEV infections for each country was calculated from the posterior FOI estimates from that country and the size of the national at-risk population. For countries where the FOI (*λ*) had been estimated from multiple studies we combined the posterior distributions of *λ* from each study into a single pooled distribution. For countries where we were unable to estimate a country-specific FOI we used a pooled distribution of the *λ* posterior distributions from all 29 studies. To capture variability in the FOI estimates the infection model was run 1000 times with random sampling from the FOI distribution. In the absence of vaccination, the number of infections at age, *a*, is calculated by multiplying the age-specific probability of infection Γ_*a*_ by the number of at-risk individuals in the age class. Estimates of age-specific population sizes were made by assuming that the age distribution of the at-risk population matched the overall national age distribution for the appropriate year from the UN World Population Prospects 2019 demographic estimates [22].

The effects of vaccination on the number of JEV infections can be estimated by adjusting the age-specific probability of infection, Γ_*a*_, to include the vaccination rate at age *a, υ*_*a*_, as in equation (4). The efficacy of a single dose of the SA 14–14–2 JEV vaccine is estimated to be 99.3% (94.9-100%) [28]. Although we estimated vaccination coverage for several study sites these estimates were not used to calculate JE burden from 2010-2019. These estimates were for the period of each study, which for some studies only partially overlaps our study period, and don’t necessarily reflect changes in vaccination coverage over the past decade. In addition, the estimates were aggregated into a handful of age classes and don’t capture within age-class variations in coverage levels. For our burden estimation process we instead used annual, age-specific vaccination coverage estimates from the literature (see SI Table 2 for references) and routine vaccination coverage estimates from WHO-UNICEF joint reporting [21].

Calculating the number of JE cases and deaths from the estimated number of JEV infections requires estimating the proportion of infections that are asymptomatic or result in only mild symptoms. The majority of human JEV infections are asymptomatic, with fewer than 1% of people infected developing clinical disease [29]. The ratio of asymptomatic infections to JE cases has been estimated as 270:1 in children aged 5-9 [30] and 300:1 for people under 40 [31], with other estimates ranging from 50:1 to 1000:1 [5]. We assume that the distribution of encephalitis cases, *C*_*a*_, observed from *I*_*a*_ JEV infections in age class *a* can be modeled with a binomial distribution *C*_*a*_ *∼ Binomial*(*I*_*a*_, *ρ*_*c*_), with a symptomatic probability of *ρ*_*c*_. The symptomatic probability, *ρ*_*c*_ was estimated by using the ‘optim’ package in R to fit a gamma distribution to the asymptomatic:symptomatic infection ratio (A:S), assuming a median A:S of 300:1 with a 95% CI of 100:1 to 750:1. The estimated gamma distribution had a shape parameter = 3.577 and a rate parameter = 0.0108. This distribution produces an A:S ratio with a median of 295:1 (95% CI: 83:1 to 717:1). The symptomatic probability was then calculated as *ρ*_*c*_ = *S/*(*S* + *A*). The mortality rate for severe encephalitis cases is believed to be between 10-40% [5]. The number of deaths from JE in age class *a, D*_*a*_, was assumed to follow *D*_*a*_ *∼Binomial*(*C*_*a*_, *ρ*_*m*_), with *ρ*_*m*_ *∼Beta*(*α* = 1.88, *β* = 3.50). This distribution produces a median mortality estimate of 0.329 (95% CI: 0.050-0.741).

Country-specific estimates of the annual number of infections, cases, and deaths were calculated by running our model 1000 times with random draws from the FOI (*λ*), symptomatic ratio (*ρ*_*c*_), and mortality rate (*ρ*_*m*_) distributions. Model simulations were started in 1950 and run through 2019 to ensure that the model approximated baseline population immunity levels from 2010 to 2019. In order to assess the impact of vaccination these estimates were generated for two scenarios: (1) using the reported vaccination coverage levels for 1950-2019, and (2) a counterfactual scenario with vaccination coverage levels set to 0 for all countries in order to estimate the impact of vaccination.

## Results

### At-risk population size estimates

Overall, 1.15 billion (range: 982.1-1543.1 million) people are estimated to live in an area suitable for endemic JEV transmission (Fig 1). This represents 28.0% (range: 24.0-37.7%) of the 4,095.5 million people living in countries with endemic JEV transmission. China had the largest number of individuals living in a likely at-risk area with 456.8 (range: 454.9-507.2) million, followed by India with 321.1 (range: 193.9-482.6) million and Bangladesh with 120.9 (range: 120.3-147.7) million (Table 1). Bangladesh had the highest percentage of its population living in at-risk areas (72.9%; range: 72.5-89.0%), followed by Vietnam (51.9%; range: 51.4-67.1%). The other countries with greater than 25% of the population living in at-risk areas were Cambodia (44.1%; range: 41.9-65.4%), Nepal (38.5%; range: 23.2-44.2%), China (32.7%; range: 32.6-36.3%), and Myanmar (32.1%; range: 30.4-43.0%). Several countries had fewer than 5% of the population living in likely at risk areas: Timor-Leste (0.5%; range: 0.2-8.5%), Bhutan (3.3%; range: 1.3-11.9%), Sri Lanka (3.9%; range: 1.3-21.6%), and Papua New Guinea (4.2%; range: 0.1-5.4%).

**Table 1.**
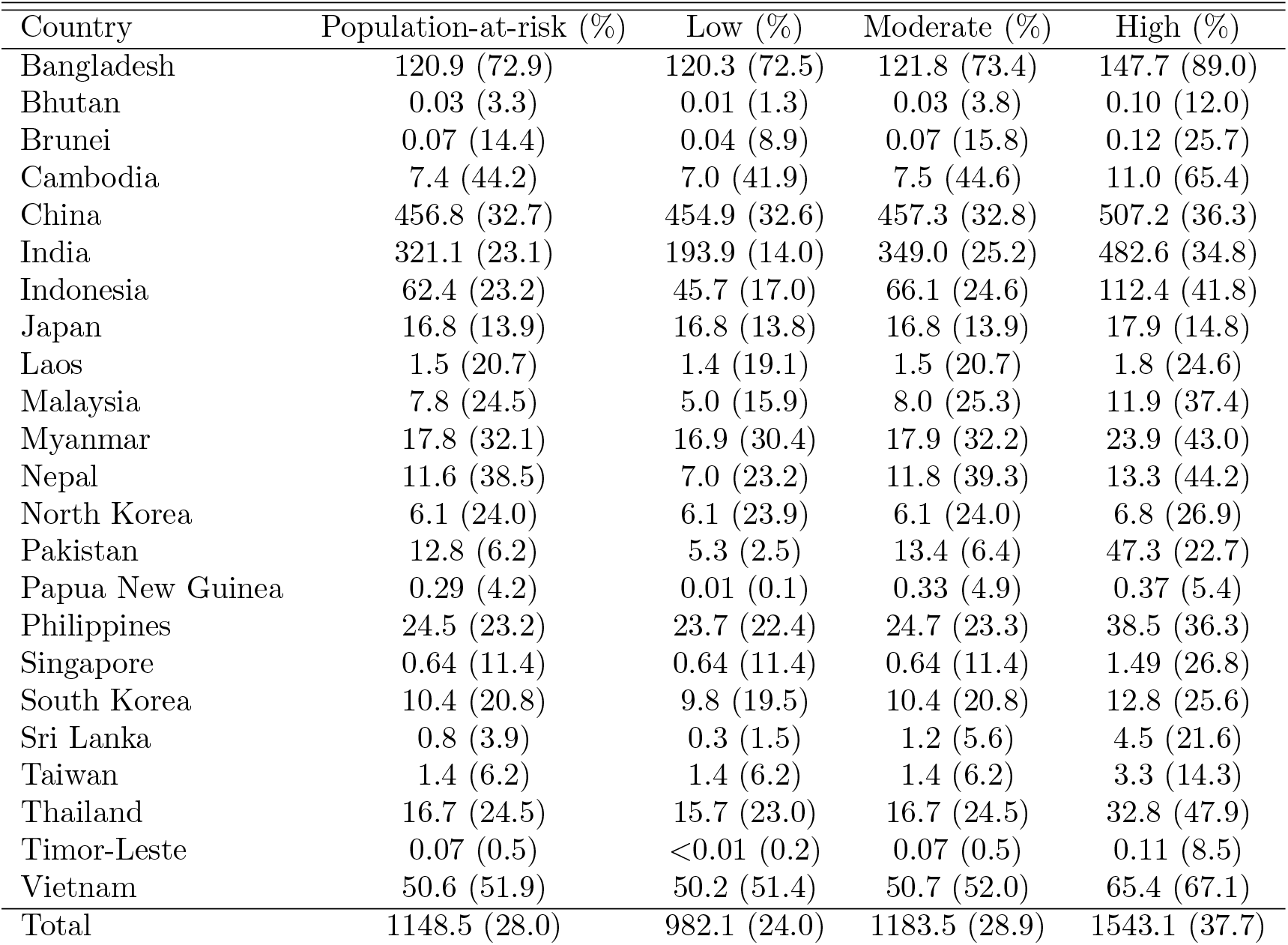
Estimates of the population at risk of JE in millions (percent of total population). Baseline estimate is for areas with wetlands or rice cultivation, suitable environmental conditions for the vector and at least 2 domestic pigs or ducks per square km. The low end estimate increases the required density of pigs or ducks to 10 per square km and the moderate estimate assumes a threshold of 1 per square km. The high estimate is for all areas classified as wetlands or rice cultivation.

| Country | Population-at-risk (%) | Low (%) | Moderate (%) | High (%) |
| --- | --- | --- | --- | --- |
| Bangladesh | 120.9 (72.9) | 120.3 (72.5) | 121.8 (73.4) | 147.7 (89.0) |
| Bhutan | 0.03 (3.3) | 0.01 (1.3) | 0.03 (3.8) | 0.10 (12.0) |
| Brunei | 0.07 (14.4) | 0.04 (8.9) | 0.07 (15.8) | 0.12 (25.7) |
| Cambodia | 7.4 (44.2) | 7.0 (41.9) | 7.5 (44.6) | 11.0 (65.4) |
| China | 456.8 (32.7) | 454.9 (32.6) | 457.3 (32.8) | 507.2 (36.3) |
| India | 321.1 (23.1) | 193.9 (14.0) | 349.0 (25.2) | 482.6 (34.8) |
| Indonesia | 62.4 (23.2) | 45.7 (17.0) | 66.1 (24.6) | 112.4 (41.8) |
| Japan | 16.8 (13.9) | 16.8 (13.8) | 16.8 (13.9) | 17.9 (14.8) |
| Laos | 1.5 (20.7) | 1.4 (19.1) | 1.5 (20.7) | 1.8 (24.6) |
| Malaysia | 7.8 (24.5) | 5.0 (15.9) | 8.0 (25.3) | 11.9 (37.4) |
| Myanmar | 17.8 (32.1) | 16.9 (30.4) | 17.9 (32.2) | 23.9 (43.0) |
| Nepal | 11.6 (38.5) | 7.0 (23.2) | 11.8 (39.3) | 13.3 (44.2) |
| North Korea | 6.1 (24.0) | 6.1 (23.9) | 6.1 (24.0) | 6.8 (26.9) |
| Pakistan | 12.8 (6.2) | 5.3 (2.5) | 13.4 (6.4) | 47.3 (22.7) |
| Papua New Guinea | 0.29 (4.2) | 0.01 (0.1) | 0.33 (4.9) | 0.37 (5.4) |
| Philippines | 24.5 (23.2) | 23.7 (22.4) | 24.7 (23.3) | 38.5 (36.3) |
| Singapore | 0.64 (11.4) | 0.64 (11.4) | 0.64 (11.4) | 1.49 (26.8) |
| South Korea | 10.4 (20.8) | 9.8 (19.5) | 10.4 (20.8) | 12.8 (25.6) |
| Sri Lanka | 0.8 (3.9) | 0.3 (1.5) | 1.2 (5.6) | 4.5 (21.6) |
| Taiwan | 1.4 (6.2) | 1.4 (6.2) | 1.4 (6.2) | 3.3 (14.3) |
| Thailand | 16.7 (24.5) | 15.7 (23.0) | 16.7 (24.5) | 32.8 (47.9) |
| Timor-Leste | 0.07 (0.5) | <0.01 (0.2) | 0.07 (0.5) | 0.11 (8.5) |
| Vietnam | 50.6 (51.9) | 50.2 (51.4) | 50.7 (52.0) | 65.4 (67.1) |
| Total | 1148.5 (28.0) | 982.1 (24.0) | 1183.5 (28.9) | 1543.1 (37.7) |

**Fig 1.**
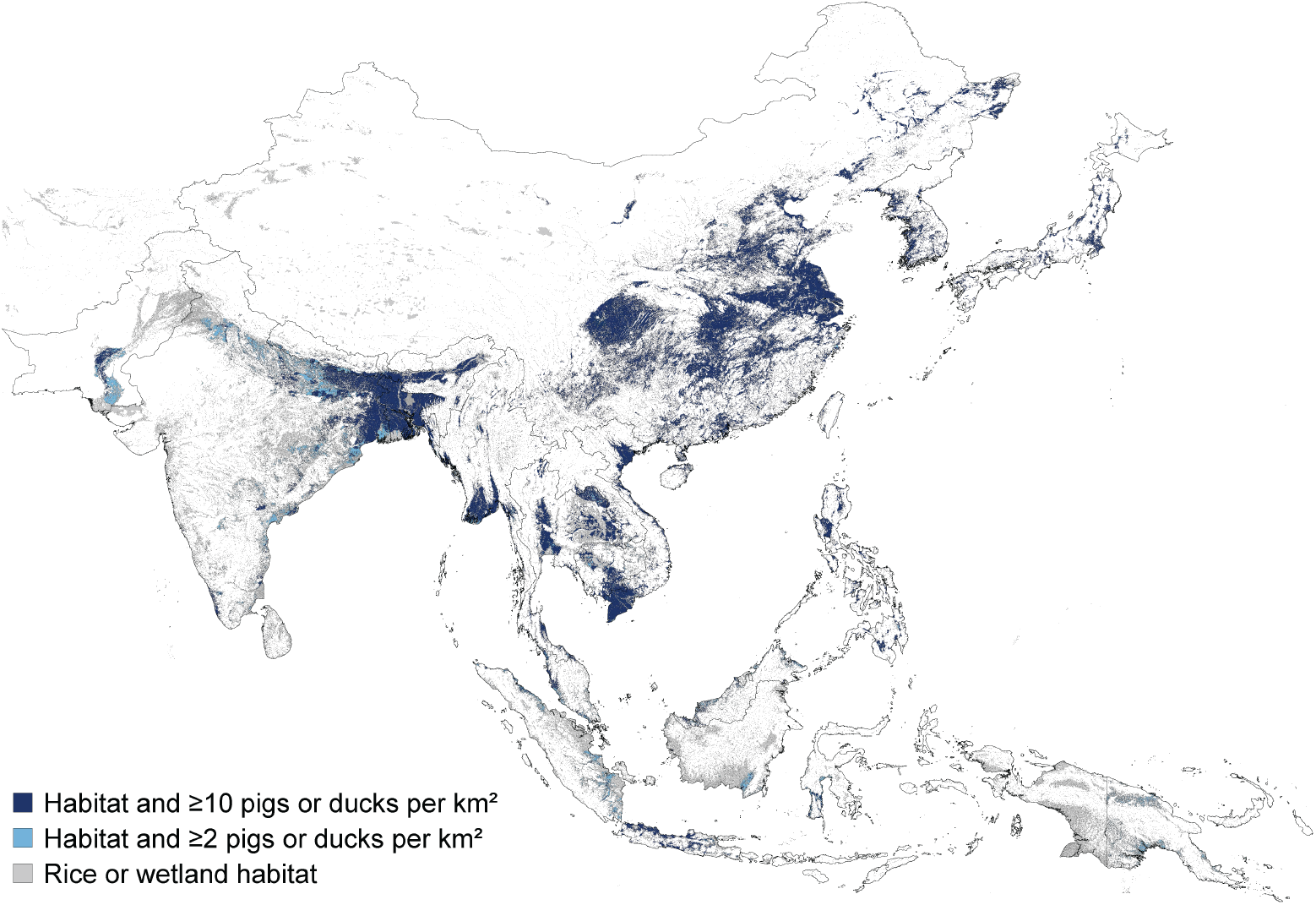
Map of the areas estimated to be suitable for endemic JEV transmission. Grey represents land areas classified as either wetland or rice cultivation and was used as the upper bound for the extent of the at-risk population. Light blue represents the default extent of the at-risk population and includes areas with wetland or rice cultivation that also have suitable environmental conditions for the mosquito vector population and a combined density of at least 2 domestic pigs and ducks per square km. Dark blue represents the lower bound on the extent of the at-risk population and only includes suitable habitat areas that have a combined density of domestic pigs and ducks of at least 10 per square km.

### Force of infection estimates

The median annual force of infection (FOI) across all studies was 0.098 (95% range: 0.012-0.354). The FOI ranged widely between countries, with median estimates for FOI ranging from a low of 0.011 (95% CrI: 0.004-0.017) in Japan to 0.286 (95% CrI: 0.125-0.437) in Indonesia (Fig 2). For countries with multiple studies, the FOI estimates from different studies often varied considerably resulting in a bimodial or more complex combined FOI distribution. Countries with significant between-study estimates in FOI included China, India, Indonesia, Malaysia, Nepal, and the Philippines (SI Table 1). Our model did not generate predicted cases that were more extreme than the numbers of observed cases—as indicated by Bayesian p-values between 0.1 and 0.9—for any of the datasets included in the analysis (SI Figs 4-30). Several potential explanatory variables were weakly associated with our FOI estimates. Higher FOI estimates were associated with less urban area (r=-0.27) and a lower per capita GDP (r=-0.27), but also lower historical incidence according to Campbell et al. [1] (r=-0.36). The best-fit model contained only two explanatory variables and had low explanatory power (BIC=1.70; adjusted *R*^2^=0.19), while the second-best model contained five explanatory variables and had a ΔBIC=0.71 (BIC=2.40; adjusted *R*^2^=0.34). Several additional models with 2 to 5 explanatory variables had ΔBIC values of <2. All models with a ΔBIC of <2 included a negative association with Campbell’s incidence classification. The best-fit model also included a negative association with the amount of land area under rice cultivation. In addition to these two explanatory variables, the second-best model included a negative association with the mean density of ducks, and a positive assocations with the mean density of chickens and the proportion of the population living in at-risk areas.

**Fig 2.**
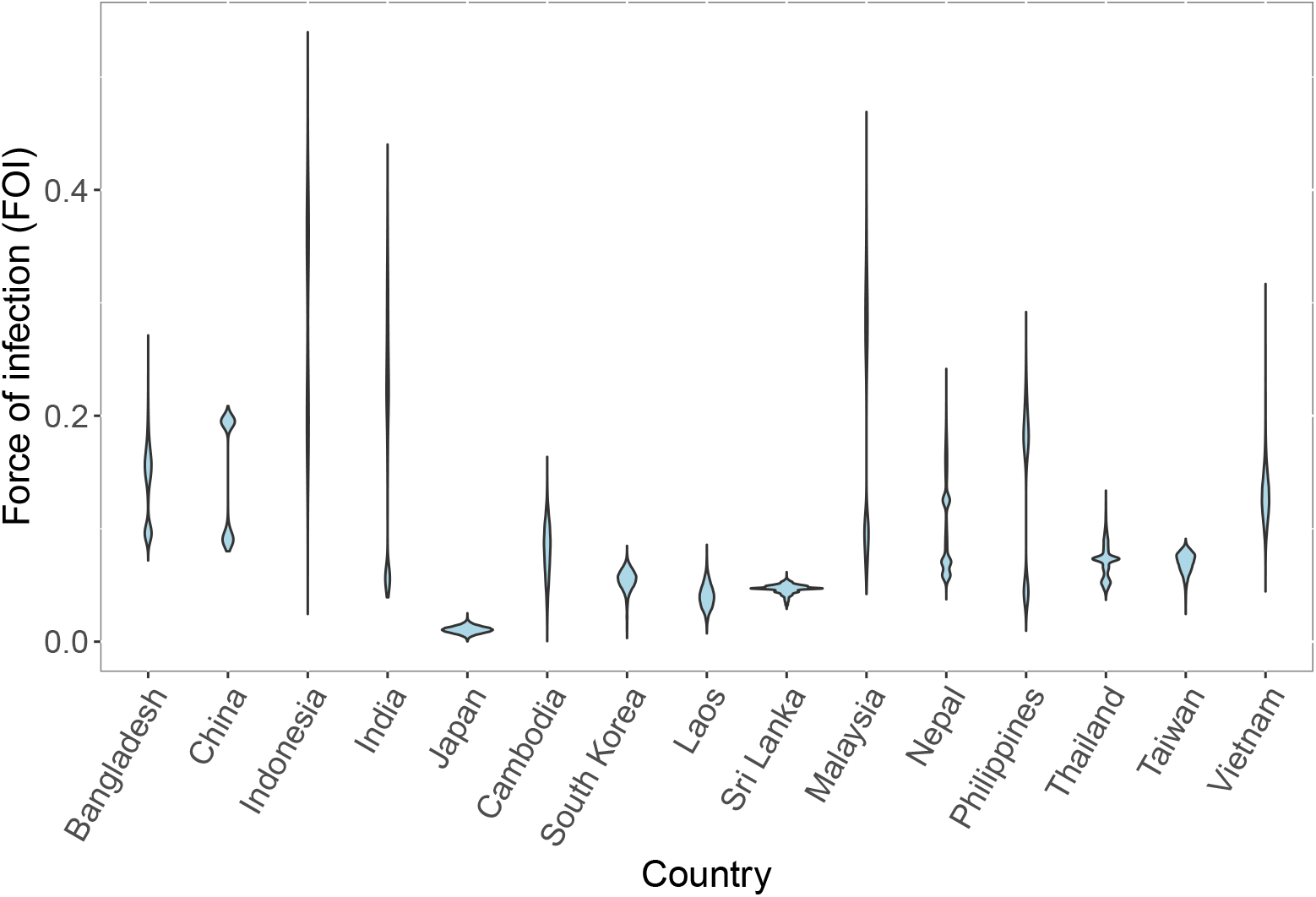
Posterior force of infection (FOI) estimates. Posterior force of infection (FOI) estimates for each country where age-specific incidence data was available. For countries where the FOI was estimated independently from more than one data source, the posterior FOI estimates for each study have been combined into a single distribution.

### Vaccination coverage estimates

The posterior estimates of vaccination coverage levels from our FOI model often differed considerably from the prior estimates derived from published studies or WHO estimates (SI Figs 31-41). In particular, the estimated vaccination coverage in the oldest age groups (typically ages 60+) was higher than the prior estimate for several studies that included older adults. These differences in vaccination coverage estimates among the oldest age groups could result from prior estimates that were too low because vaccinations that occurred several decades ago were missing from the data sources we used. Our model would also overestimate vaccination coverage in older individuals if transmission intensity was higher in the past. The posterior estimates of vaccination coverage in younger children were lower than our prior estimates for studies conducted in Japan and Malaysia (SI Figs 35,38). In India, South Korea, and Taiwan vaccination coverage estimates in younger children were higher than posterior estimates (SI Figs 33-40).

### Estimates of Japanese encephalitis burden and vaccination impacts

Based on the baseline number of people at risk of infection, the mean number of JE cases decreased from 61,988 (95% CI: 19,510-210,105) in 2010 to 43,683 (95% CI: 13,750-142,239) in 2019 (Fig 3A). The mean number of JE deaths decreased from 22,522 (95% CI: 2,467-86,068) in 2010 to 15,861 (95% CI: 1,754-59,169) in 2019. In the absence of any vaccination we estimate there would have been 74,739 (95% CI: 23,256-246,636) cases and 27,179 (95% CI: 2,964-103,521) deaths in 2019. Between 2010 and 2019, vaccination prevented 229,629 (95% CI: 69,648-759,615) cases and 83,849 (95% CI: 8,551-320,853) deaths. India had the largest estimated JE burden in 2019 with 17,744 (95% CI: 5,638-56,911) cases and 6,451 (95% CI: 701-24,290) deaths (Fig 4). The next highest estimated burdens were in Bangladesh and China with 9,274 (95% CI: 2,909-30,646) and 5,575 (95% CI: 910-20,217) cases and 3,375 (95% CI: 382-12,921) and 1,996 (95% CI: 145-8,077) deaths respectively (Fig 4). In the absence of vaccination we estimate that China, rather than India, would have had the highest JE burden in 2019 with 24,584 (95% CI: 7,668-79,936) cases and 8,937 (95% CI: 973-33,675) deaths. From 2010-2019, we estimate that vaccination had the largest impact in China, with 148,408 (95% CI: 53,791-483,052) cases and 54,289 (95% CI: 6,382-208,611) deaths prevented. Taiwan and Malaysia had the largest percent reductions in JE burden due to vaccination, with 91.1% and 80.5% reductions in JE cases respectively.

**Fig 3.**
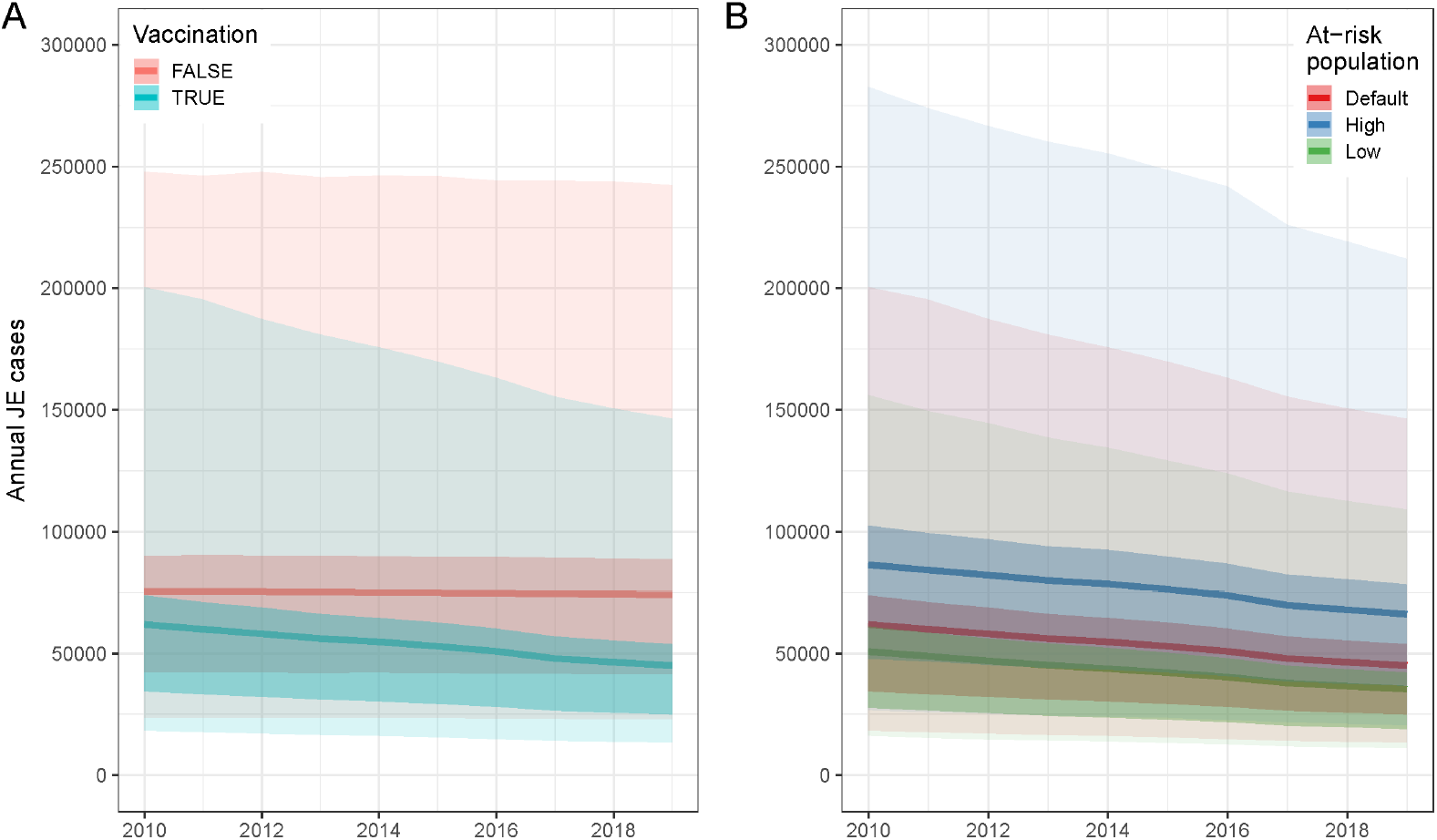
Annual Japanese encephalitis (JE) cases from 2010-2019. (A) Annual JE cases from 2010 to 2019 with vaccination (blue) or under a counterfactual scenario where vaccination coverage in all countries was zero (red). Estimates based on default at-risk population size. (B) Estimates of annual JE cases from 2010 to 2019 using three different estimates of the total size of the at-risk population. Darker bands represents the interquartile range (IQR) and lighter bands represents the 95% confidence intervals (CIs).

**Fig 4.**
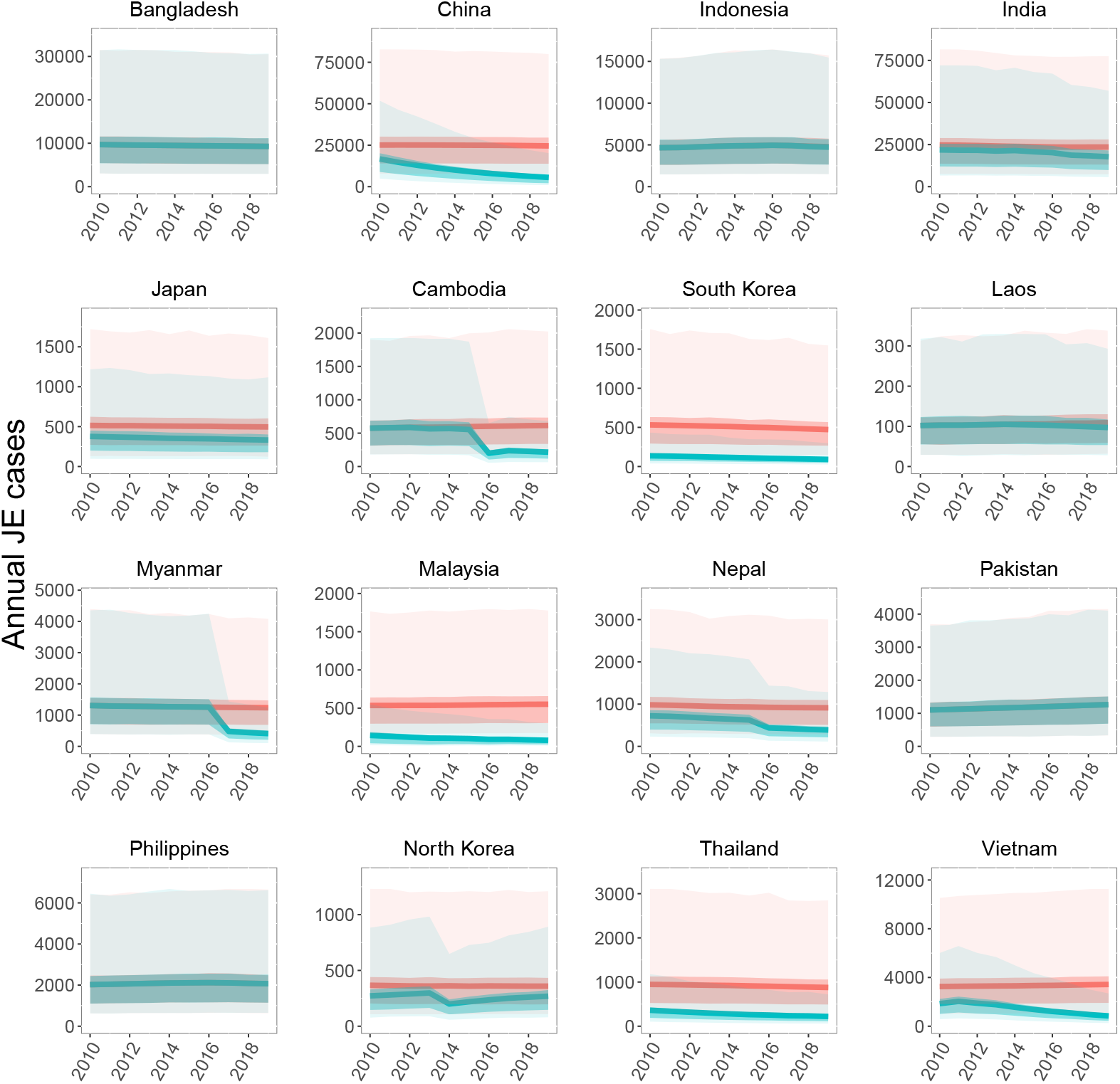
National-level Japanese encephalitis (JE) cases from 2010-2019. Annual national-level JE cases from 2010 to 2019 with vaccination (blue) or under a counterfactual scenario where vaccination coverage in all countries was zero (red). Displayed results are limited to the 16 countries with more than 100 mean annual cases in the absence of vaccination. Darker bands represents the interquartile range (IQR) and lighter bands represents the 95% confidence intervals (CIs).

Using the lower estimate for the size of the population at risk of JEV infection leads to an estimated 34,211 (95% CI: 10,706-109,739) cases and 12,414 (95% CI: 1,368-47,386) deaths in 2019 compared to the estimate of 43,683 (95% CI: 13,750-142,239) cases and 15,861 (95% CI: 1,754-59,169) deaths using our default estimate for the size of the at-risk population (Fig 3B). With the highest estimate for the size of the at-risk population, we estimate a mean total of 64,645 (95% CI: 20,550-208,745) cases and 23,480 (95% CI: 2,627-89,382) deaths in 2019. India and Bangladesh remained the countries with the highest and second-highest estimated JE burdens in 2019 under all at-risk population size estimates; however, with the high estimate of the at-risk population size, the country with the third highest burden is Indonesia rather than China (SI Table 3).

## Discussion

Despite recent increases in vaccination coverage, Japanese encephalitis (JE) remains a major cause of morbidity and mortality throughout South and East Asia. Based on an examination of the environmental factors and land use characteristics associated with JEV transmission, we estimate that over one billion people live in areas suitable for endemic transmission of JEV. The JE burden in endemic countries decreased from approximately 62,000 cases and 22,500 deaths in 2010 to fewer than 44,000 cases and 16,000 deaths in 2019, mainly due to increases in vaccination coverage over the past decade. We estimate that vaccination prevented over almost 230,000 cases and 84,000 deaths during the 2010s. Our estimates highlight countries that have significantly reduced their burden through vaccination in recent years (e.g., China, Cambodia, and Vietnam), as well as countries where the burden remains high and increasing vaccination coverage could have a large impact (e.g., Bangladesh, India, and the Philippines).

The estimated at-risk population of 1.15 (range: 0.98-1.54) billion in potentially JEV-endemic areas of Asia represents 28.0% (range: 24.0-37.7%) of the total 2020 population of just over four billion in these endemic countries. The lower end of the estimated range represents the population size in areas where the local habitat type and climate are suitable for JEV transmission and suitable reservoir hosts are present at a high density, while the upper end of the range requires only a suitable habitat type and does not consider the known distributions of the vector or reservoir species. Therefore, we estimate that over 1.5 billion people within the endemic region of Asia live in an area with land that is either used for rice cultivation [9] or within several kilometers of wetlands (including lakes, rivers, or streams). When we further restricted the at-risk population to areas within the predicted distribution of the main vector species *C. tritaeniorhynchus* [12] or with a warm, wet climate suitable for other potential vector species, and with an estimated density of at least two domestic pigs or ducks per square km, the estimated size of the at-risk population was 1.15 billion. Lowering the pig and duck density threshold to one per square km slightly raised the at-risk population to 1.18 billion, while raising the threshold to ten per square km lowered the size of the at-risk population to just under one billion (982 million). Although non-human hosts are a requisite part of the transmission cycle, the extent to which different domestic livestock species serve as reservoir or amplifying hosts is still uncertain [3]. As a result, the different livestock density thresholds that we considered in our analysis are only preliminary assumptions, and further study of the importance of different wildlife and domestic species to spillover is required. However, our results were not overly sensitive to these assumptions as ranging the threshold from one to ten animals per square km only varied the size of the at-risk population from 0.98 to 1.18 billion.

Our estimate of the size of the at-risk population represents a significant reduction compared to a previous estimate used to calculate the global incidence of JE in 2010 [1]. Campbell et al. [1] reported that 3.15 billion people lived in JE endemic areas in 2010 out of a total population of 3.69 billion in endemic and non-endemic areas in the region. However, they classified all of China as either historically medium- or high-incidence endemic areas, while we estimate that only 32.7% (range: 32.6-36.3%) of the Chinese population lives in JE-endemic areas. Over the entire region, Campbell et al. [1] only excluded portions of a few countries—such as India, Pakistan, and Nepal—as non-endemic areas. In contrast, even using our high-end estimates, the portion of the at-risk population is only higher than 50% in Bangladesh (89.0%), Vietnam (67.1%), and Cambodia (65.4%). The upper estimates of the at-risk populations of Thailand, Nepal, Myanmar, and Indonesia are between 40 and 50%, while the remaining countries are all below 40%. We estimate that only a small percentage (<15%) of the population in several endemic countries—Bhutan, East Timor, and Papua New Guinea—lives in likely JE-endemic areas. We also estimate that several countries—including Singapore, Japan, South Korea, and Taiwan—that used to have high JE incidence rates now have relatively small at-risk populations (even before vaccination) due to dramatic shifts away from rural agriculture via urbanization and industrialization over the past 50+ years. One additional difference is that Campbell et al. [1] also included portions of Australia and Russia as historically endemic regions, while we excluded these two countries because Russia has not reported a JE case in over a decade and Australia has only reported a handful of locally-acquired cases on several islands in the Torres Strait and a small region of Cape York in northern Queensland [32].

Although our estimate of the size of the at-risk population is considerably smaller than the 3.15 billion people living in JE endemic areas according to Campbell et al. ([1]), our estimate of 61,988 (95% CI: 19,510-210,105) cases in 2010 is similar to their estimate of approximately 67,900 cases. We estimated a higher per capita incidence rate in many countries compared to Campbell et al. [1] based on our FOI estimates derived from age-specific case data. In contrast, Campbell et al. [1] extrapolated global incidence from per capita incidence rates in several different regions (classified by both historical incidence and the current level of vaccination in the region), hence their per capita incidence rates were based on a larger fraction of the population in a region than our at-risk population estimates. A recent study by Quan et al. [20] estimated that there were over 100,000 (95% CI: 61,720-157,522) JE cases in 2015, an estimate over 50% higher than our estimate (although there is overlap in the 95% confidence intervals of the two estimates). Quan et al. [20] used a similar method of first estimating the FOI in each country based on age-specific case data obtained from a systematic review. These country-specific FOI estimates were then used to estimate incidence in each country using the regional classifications and endemic population sizes from Campbell et al. [1]. This combination of high FOIs (in some countries) and a larger at-risk population leads to the higher incidence estimates of Quan et al. [20] relative to our estimates and those of Campbell et al. [1]. As a result, for several countries, such as Bangladesh and Pakistan, where we estimate that the at-risk population is a fairly large percentage of the population in the “endemic” region of the country, our incidence estimates are similar to the estimates of Quan et al. [20]. For several countries—including Laos, Malaysia, Myanmar, Philippines, and Thailand—the incidence estimates of Quan et al. [20] are two to four times higher than our estimates due to our lower estimates of the sizes of the at-risk populations. These discrepancies in estimated incidence rates and the overall burden of JE arising from different assumptions about how many people are at risk of infection emphasizes the importance of refining our understanding of JE epidemiology and endemicity. Particularly important questions are (a) the extent to which JEV transmission and spillover to humans occurs in areas where rice cultivation is limited or non-existent, and (b) to what extent domestic animals other than pigs serve as reservoir or amplifying hosts.

We estimated that vaccination prevented over 229,000 cases and 83,000 deaths between 2010 and 2019. This a similar annual impact to the 308,000 cases and 75,000 prevented by vaccination between 2000 and 2015 as estimated by Quan et al. ([20]). However, as overall vaccination coverage has increased over time, our vaccination impact estimates for the overlapping period of the two studies (2010-2015) is lower due to our lower incidence estimates. Although the absolute impact of vaccination is lower in our estimation, the impact is similar when expressed as a percentage of cases prevented. Quan et al. ([20]) estimated that vaccination reduced JE incidence in 2015 from 145,542 (95% CI: 96,667–195,639) to 100,308 (95% CI: 61,720–157,522), a 31.1% reduction, while we estimate that vaccination reduced 2015 incidence from 75,635 (95% CI: 23,585-250,449) to 52,286 (95% CI: 16,524-170,911), a 30.9% reduction. Our estimate of the impact of vaccination on JE burden over the past decade is likely somewhat conservative because information on some sub-national vaccination campaigns is missing from the datasets we obtained.

From 2010 to 2019, the largest absolute impact of vaccination occurred in China, where the JE vaccine was added to the national routine vaccination program for infants in 2008 [33]. We also estimate that increased vaccination coverage in 2015 or 2016 in Cambodia, Myanmar, and Nepal has led to substantial reductions in incidence over the past few years in these countries. Our estimates also indicate that incidence remains relatively low in the countries—Japan, Malaysia, South Korea, Sri Lanka, Taiwan, and Thailand—where vaccination has been common for over a decade. In addition, Vietnam, which began wide-scale vaccination in 1997, prevented an estimated 18,759 (95% CI: 5,960-62,711) cases and 6,818 (95% CI: 778-26,012) deaths between 2010 and 2019, resulting in a 54.4% decline in incidence over that time period. However, due to the large size of the at-risk population and the high FOI in Vietnam, we estimate that there were still almost one thousand (95% CI: 244-2,685) JE cases in 2019. Our vaccination impact estimates also highlight the countries where vaccine introduction or an increase in vaccination coverage could have the largest impact. Bangladesh, Indonesia, Pakistan, and Philippines all have over one thousand estimated JE cases annually and do not have national vaccination programs. While vaccination coverage has increased in India over the past decade, we estimate that there were still over 17,700 (95% CI: 5,638-56,911) cases and 6,400 (95% CI: 701-24,290) deaths in 2019. In addition to adding the JE vaccine to the routine vaccination schedule, in order to rapidly reduce their JE burden countries should also consider vaccination campaigns to cover older children (and potentially adults) in endemic areas. Estimates of the FOI in each country could be helpful for determining the appropriate age group to target, as the FOI determines the proportion of older children and adults that still remain susceptible to infection [8].

We estimate that transmission intensity is relatively high in many JE-endemic countries. The median FOI across all studies was 0.098, which corresponds to an 9.3% annual probability of a susceptible individual living in an at-risk area being infected. Several countries with large at-risk populations—including Bangladesh, China, India, Indonesia, Philippines, and Vietnam—had higher than average FOI estimates. At the national level we found that there was a negative relationship between FOI and the proportional extent of urban areas, which aligns with the current understanding of JE as a disease that mainly affects rural areas [34]. FOI estimates were also higher for countries with lower per-capita GDPs, suggesting that economic development (often coupled with urbanization) can reduce transmission intensity by lowering contact rates between humans and JEV vectors and reservoir hosts. Overall, we did not find any strong correlations between our FOI estimates and the potential explanatory variables and the best-fit regression model explained little of the variance in the FOI estimates. Any correlations between our FOI estimates and national-level averages for different covariates should be viewed with caution as our analysis is based on fewer than 30 studies and national-level averages can obscure important within-country heterogeneities. In addition, there were some moderate to strong correlations between covariates that complicate the relationships between these covariates and the FOI. For example, the mean density of pigs was moderately correlated with both the proportional urban area of a country (r=0.65) and national GDP (r=0.57), which may explain why we found a weakly negative correlation between FOI and the mean density of pigs (r=-0.20) despite the role of pigs to human spillover. In addition, in the countries we studied the average extent of rice cultivation was highly correlated with higher mean densities of chickens, ducks, or cows (r*>*0.75), but not pigs.

Our analysis of the at-risk population size has several limitations. We restricted the suitable habitat for JEV transmission to areas with rice cultivation or adjacent wetlands, however, the large-scale wetlands dataset does not capture smaller pockets of potentially suitable habitat such as small water courses, ponds, or artificial reservoirs that could serve as vector habitat. In addition, the extent to which lands that are irrigated for crops other than rice can support mosquito populations that transmit JEV is unknown. Both of these potential sources of additional vector habitat could increase our estimate of the at-risk population size. Our estimates are also limited by current gaps in the knowledge of the ecologies of the various JEV vector species [5]. We used a modeled distribution for the primary vector, *C. tritaeniorhynchus*, which may not be accurate in certain areas [12]. Many other mosquito species have been shown to be competent vectors in lab studies or have been documented as JEV-positive in field studies [5]. However, our knowledge of the distribution of these vector species is limited, as is the extent to which they are capable of maintaining JE endemicity or serving as bridge vectors to humans in the absence of *C. tritaeniorhynchus*. Additional field studies are required to assess the role of these alternative vector species, but low rates of viral positivity in mosquito populations often make it difficult to determine their importance in the transmission cycle [35, 36]. A further source of uncertainty in our at-risk estimates arises from the uncertainty surrounding the extent to which different species can serve as reservoir and amplifying hosts in the JEV transmission cycle. While domestic pigs have been associated with spillover to humans [3], endemic transmission has also been documented in areas where there are few or no pigs, suggesting that other species are involved in the transmission cycle and spillover in these areas [37, 38]. Domesticated waterfowl and chickens are capable of infection, but evidence for their roles as reservoir or amplifying hosts is still weak [3, 37, 39, 40]. In some areas, wild waterfowl may be abundant enough to sustain endemic transmission and occasional spillover to humans [5]. Field studies in endemic areas where pigs are absent could help clarify the role of these other species in the transmission cycle and identify key factors associated with spillover to humans.

In addition to uncertainties regarding the size of the at-risk population, our JE burden estimates have several additional limitations. One limitation is that we have assumed that there is no sub-national variation in the FOI. The differences in FOI estimates for several countries where we were able to estimate FOI independently from multiple studies indicates that there may be substantial sub-national variation. Where available, we have used the pooled posterior FOI estimates from multiple studies to try and capture the possible range, but this option was not available for all countries. Further studies are needed to determine the extent of sub-national variation in transmission intensity and to identify the factors associated with this variation. We have also assumed that the FOI remained constant over time due to the limited amount of age-specific incidence data available to derive time-varying FOI estimates. However, it is likely that changes in land use, agriculture practices, and development have affected transmission intensity in complex ways throughout Asia [2]. For example, we estimate that the FOI is low in several countries (e.g., Japan, Taiwan, and South Korea) that had high incidence rates over 50 years ago, but have since experienced large-scale economic development and population shifts away from small, rural farms. In contrast, the increased proximity of pig rearing and rice cultivation in other areas may have increased transmission intensity in some countries [2]. We also were not able to estimate an FOI for several countries due to a lack of age-specific incidence data. For these countries we sampled from the full distribution of FOI values seen in our estimates, which increases the uncertainty regarding the current JE burden for these countries. Improved surveillance or epidemiological studies in these countries would help refine these estimates.

## Conclusion

By estimating the FOI experienced per susceptible individual in different JE-endemic countries, we were able to estimate the total number of JE cases and deaths under different vaccination scenarios over the past decade. By coupling these FOI estimates with estimates of the size of the at-risk population from a spatial analysis of the risk factors associated with JEV transmission, we were able to estimate the JE burden even for countries where JE is currently under-reported. Comparing our estimates of at-risk population sizes based on knowledge of the environmental factors associated with JE endemicity to a prior estimate of the population living in JE-endemic regions [1] highlights the importance of refining our understanding of where JE is endemic and spillover to humans occurs regularly. Understanding the sub-national variation in infection risk will be key to targeting vaccination campaigns towards the most vulnerable populations. By modeling current JE incidence against a counterfactual scenario of incidence in the absence of vaccination, we estimated the progress each country has made towards reducing their JE burden through vaccination over the past decade. We have also identified which countries could benefit the most from the introduction or expansion of vaccination coverage.

## Data Availability

All data used in our analysis is publicly available through the data references described in the manuscript.

https://github.com/mooresea/JEV_estimation

## Acknowledgements

We thank the members of the Perkins lab at the University of Notre Dame for reviewing this manuscript. We also thank several members of the Vaccine Impact Modelling Consortium for reviewing and recommending improvements to the methodology used in this analysis.

## Supporting Information

**Table S1. Summary information and results for studies of age-specific Japanese encephalitis (JE) incidence used to estimate the annual force of infection (FOI)**.

**Table S2. Summary of vaccination coverage data used to estimate the burden of Japanese Encephalitis (JE) from 2010-2019 for each country**.

**Table S3. Mean annual number of JE cases (C) and deaths (D) per country from 2010 to 2019 under the vaccination and no vaccination scenarios**.

**S1 Fig. Spatial extent of rice cultivation and wetlands in Asia**.

**S2 Fig. Map of likely suitable habitat for JEV vector species**.

**S3 Fig. Combined population densities per kilometer of domestic pigs and ducks**.

**S4 Fig. Observed versus predicted number of JE cases per age class in Bangladesh based on dataset from [15]**.

**S5 Fig. Observed versus predicted number of JE cases per age class in Cambodia based on dataset from [41]**.

**S6 Fig. Observed versus predicted number of JE cases per age class in Cambodia based on dataset from [42]**.

**S7 Fig. Observed versus predicted number of JE cases per age class in China based on dataset from [43]**.

**S8 Fig. Observed versus predicted number of JE cases per age class in China based on dataset from [44]**.

**S9 Fig. Observed versus predicted number of JE cases per age class in India based on dataset from [45]**.

**S10 Fig. Observed versus predicted number of JE cases per age class in India based on dataset from [46]**.

**S11 Fig. Observed versus predicted number of JE cases per age class in India based on dataset from [47]**.

**S12 Fig. Observed versus predicted number of JE cases per age class in Indonesia based on dataset from [48]**.

**S13 Fig. Observed versus predicted number of JE cases per age class in Indonesia based on dataset from [16]**.

**S14 Fig. Observed versus predicted number of JE cases per age class in Japan based on dataset from [49]**.

**S15 Fig. Observed versus predicted number of JE cases per age class in Laos based on dataset from [50]**.

**S16 Fig. Observed versus predicted number of JE cases per age class in Malaysia based on dataset from [51]**.

**S17 Fig. Observed versus predicted number of JE cases per age class in Malaysia based on dataset from [19]**.

**S18 Fig. Observed versus predicted number of JE cases per age class in Nepal based on dataset from [52]**.

**S19 Fig. Observed versus predicted number of JE cases per age class in Nepal based on dataset from [53]**.

**S20 Fig. Observed versus predicted number of JE cases per age class in Nepal based on dataset from [54]**.

**S21 Fig. Observed versus predicted number of JE cases per age class in Nepal based on dataset from [55]**.

**S22 Fig. Observed versus predicted number of JE cases per age class in Nepal based on dataset from [18]**.

**S23 Fig. Observed versus predicted number of JE cases per age class in the Philippines based on dataset from [56]**.

**S24 Fig. Observed versus predicted number of JE cases per age class in the Philippines based on dataset from [57]**.

**S25 Fig. Observed versus predicted number of JE cases per age class in the Philippines based on dataset from [58]**.

**S26 Fig. Observed versus predicted number of JE cases per age class in South Korea based on dataset from [59]**.

**S27 Fig. Observed versus predicted number of JE cases per age class in Sri Lanka based on dataset from [60]**.

**S28 Fig. Observed versus predicted number of JE cases per age class in Taiwan based on dataset from [61]**.

**S29 Fig. Observed versus predicted number of JE cases per age class in Thailand based on dataset from [62]**.

**S30 Fig. Observed versus predicted number of JE cases per age class in Vietnam based on dataset from [63]**.

**S31 Fig. Estimated vaccination coverage by age group for China based on age-specific incidence data from Li et al. [43]**.

**S32 Fig. Estimated vaccination coverage by age group for China based on age-specific incidence data from Wang et al. [44]**.

**S33 Fig. Estimated vaccination coverage by age group for India based on age-specific incidence data from Ranjan et al. [45]**.

**S34 Fig. Estimated vaccination coverage by age group for India based on age-specific incidence data from Jain et al. [47]**.

**S35 Fig. Estimated vaccination coverage by age group for Japan based on age-specific incidence data from Arai et al. [49]**.

**S36 Fig. Estimated vaccination coverage by age group for South Korea based on age-specific incidence data from Lee et al. [59]**.

**S37 Fig. Estimated vaccination coverage by age group for Sri Lanka based on age-specific incidence data from the Epidemiology Unit of the Sri Lankan Ministry of Health [60]**.

**S38 Fig. Estimated vaccination coverage by age group for Malaysia based on age-specific incidence data from Mustapa et al. [51]**.

**S39 Fig. Estimated vaccination coverage by age group for Thailand based on age-specific incidence data from Olsen et al. [62]**.

**S40 Fig. Estimated vaccination coverage by age group for Taiwan based on age-specific incidence data from Chang et al. [61]**.

**S41 Fig. Estimated vaccination coverage by age group for Vietnam based on age-specific incidence data from Yen et al. [63]**.

## Notes

### Competing Interest Statement

The authors have declared no competing interest.

### Funding Statement

This work was carried out as part of the Vaccine Impact Modelling Consortium (www.vaccineimpact.org), which is funded by Gavi, the Vaccine Alliance, and the Bill & Melinda Gates Foundation (OPP1157270). The views expressed are those of the authors and not necessarily those of the Consortium or its funders. The funders were given the opportunity to review this paper prior to publication, but the final decision on the content of the publication was taken by the authors. The funders had no role in study design, data collection and interpretation, or the decision to submit the work for publication.

