## Supplementary Materials for "The current burden of Japanese encephalitis and the estimated impacts of vaccination: Combining estimates of the spatial distribution and transmission intensity of a zoonotic pathogen"

### Supporting Information

**Table S1. Summary information and results for studies of age-specific Japanese encephalitis (JE) incidence used to estimate the annual force of infection (FOI).** Results are the median posterior FOI estimates with 95% credible interval. Vaccination coverage was assumed to be 0 for studies without a vaccination data source.

| Country | Study period | FOI (95% CrI) | Incidence data | Vaccination data |
|---|---|---|---|---|
| Bangladesh | 2007-2008 | 0.148 (0.087 - 0.197) | Paul et al. (2011) | |
| Cambodia | 2006-2008 | 0.067 (0.027 - 0.107) | Touch et al. (2009) | |
| Cambodia | 2010-2013 | 0.096 (0.064 - 0.129) | Horwood et al. (2017) | |
| China | 2004-2014 | 0.091 (0.085 - 0.104) | Li et al. (2016) | 2000-2014: WHO (2020) |
| China | 2000-2005 | 0.195 (0.193 - 0.197) | Wang et al. (2009) | 2000-2005: WHO (2020) |
| India | 2008-2012 | 0.281 (0.150 - 0.372) | Ranjan et al. (2014) | 2006-2012: WHO (2020) |
| India | 2000-2002 | 0.056 (0.046 - 0.066) | Phukan et al. (2004) | |
| India | 2011-2013 | 0.224 (0.159 - 0.295) | Jain et al. (2016) | 2006-2013: WHO (2020) |
| Indonesia | 2001-2003 | 0.197 (0.112 - 0.283) | Kari et al. (2006) | |
| Indonesia | 2005-2006 | 0.365 (0.287 - 0.451) | Ompusunggu et al. (2008) | |
| Japan | 1982-2004 | 0.011 (0.004 - 0.017) | Arai et al. (2008) | 1976-2004: Quan et al. (2020) |
| Laos | 2001-2008 | 0.040 (0.021 - 0.062) | Moore et al. (2012) | |
| Malaysia | 2006-2013 | 0.094 (0.060 - 0.117) | Mustapa et al. (2016) | 2001-2013: WHO (2020) |
| Malaysia | 1997-2001 | 0.290 (0.230 - 0.375) | Wong et al. (2008) | |
| Nepal | 2004-2006 | 0.059 (0.055 - 0.063) | Wierzba et al. (2008) | |
| Nepal | 1996-1997 | 0.071 (0.068 - 0.074) | Joshi (2007) | |
| Nepal | 2006 | 0.084 (0.057 - 0.117) | Partridge et al. (2007) | |
| Nepal | 2007 | 0.161 (0.129 - 0.198) | Bhattachan et al. (2009) | |
| Nepal | 2004-2006 | 0.125 (0.119 - 0.131) | Pant (2009) | |
| Philippines | 2011-2014 | 0.178 (0.153 - 0.205) | Lopez et al. (2015) | |
| Philippines | 2002-2004 | 0.044 (0.028 - 0.064) | Nativad et al. (2006) | |
| Philippines | 1984 | 0.200 (0.162 - 0.244) | Luis et al. (1990) | |
| South Korea | 2001-2014 | 0.055 (0.032 - 0.071) | Lee et al. (2016) | 1963-1999: Sohn (2000), Monath (2002)<br>2000-2005: Quan et al. (2020)<br>2006-2014: WHO (2020) |
| Sri Lanka | 2006-2010 | 0.047 (0.035 - 0.054) | Epidemiology Unit MoH (2012) | 1988-2008: Quan et al. (2020)<br>2008-2010: Gavi |
| Taiwan | 2000-2014 | 0.071 (0.049 - 0.084) | Chang et al. (2017) | 1969-2014: Monath (2002), Hsu et al. (2014) |
| Thailand | 2003-2005 | 0.073 (0.046 - 0.102) | Olsen et al. (2010) | 1990-2005: Olsen et al. (2010), Quan et al. (2020) |
| Vietnam | 1998-2007 | 0.128 (0.088 - 0.185) | Yen et al. (2010) | 1997-2005: Choisy (2017)<br>2006-2007: WHO (2020) |

**Table S2. Summary of vaccination coverage data used to estimate the burden of Japanese encephalitis (JE) from 2010-2019 for each country.** Countries without details were assumed to have a vaccination coverage of 0.

| Country | Start | End | Source |
|---|---|---|---|
| Bangladesh | | | |
| Bhutan | | | |
| Brunei | | | |
| Cambodia | 2013 | 2019 | WHO (2016), WHO (2020), Quan et al. (2020) |
| China | 2008 | 2019 | WHO (2020) |
| East Timor | | | |
| India | 2006 | 2019 | WHO (2020), Quan et al. (2020) |
| Indonesia | 2018 | 2019 | Im et al. (2018), WHO (2020) |
| Japan | 1976 | 2006 | Quan et al. (2020) |
| | 2007 | 2019 | WHO (2020) |
| Laos | 2015 | 2016 | MoH Laos (2016) |
| Malaysia | 2001 | 2019 | WHO (2020) |
| Myanmar | 2017 | 2019 | WHO (2018), WHO (2020) |
| Nepal | 2005 | 2019 | MoH Nepal (2016), Mithia Public Health Services, Nepal (2016), WHO (2020) |
| North Korea | 2009 | 2014 | Marks et al. (2015) |
| Pakistan | | | |
| Papua New Guinea | | | |
| Philippines | | | |
| Singapore | | | |
| South Korea | 1963 | 1999 | Sohn (2000), Monath (2002) |
| | 2000 | 2005 | Quan et al. (2020) |
| | 2006 | 2019 | WHO (2020) |
| Sri Lanka | 1988 | 2008 | Quan et al. (2020) |
| | 2009 | 2019 | WHO (2020) |
| Taiwan | 1968 | 2019 | Monath (2002), Hsu et al. (2014) |
| Thailand | 1990 | 2015 | Olsen et al. (2010), Quan et al. (2020) |
| | 2016 | 2019 | WHO (2020) |
| Vietnam | 1997 | 2005 | Choisy (2017) |
| | 2006 | 2019 | WHO (2020) |

Ministry of Health, Laos. 2016. Japanese encephalitis SIA 2015 technical report.

Ministry of Health, Nepal. 2016. Japanese encephalitis vaccination campaign, Nepal. Technical Report. WHO-UNICEF Joint Reporting.

Mithila Public Health Services, Nepal. 2016. Coverage survey of Japanese encephalitis vaccine mass vaccination campaign in selected districts of Nepal.

Monath, T.P., 2002. Japanese Encephalitis Vaccines: Current Vaccines and Future Prospects, in: Mackenzie, J.S., Barrett, A.D.T., Deubel, V. (Eds.), Japanese Encephalitis and West Nile Viruses, Current Topics in Microbiology and Immunology. Springer, Berlin, Heidelberg, pp. 105–138. https://doi.org/10.1007/978-3-642-59403-8_6

Olsen, S.J., Supawat, K., Campbell, A.P., Anantapreecha, S., Liamsuwan, S., Tunlayadechanont, S., Visudtibhan, A., Lupthikulthum, S., Dhiravibulya, K., Viriyavejakul, A., Vasiknanonte, P., Rajborirug, K., Watanaveeradej, V., Nabangchang, C., Laven, J., Kosoy, O., Panella, A., Ellis, C., Henchaichon, S., Khetsuriani, N., Powers, A.M., Dowell, S.F., Fischer, M., 2010. Japanese encephalitis virus remains an important cause of encephalitis in Thailand. International Journal of Infectious Diseases 14, e888–e892. https://doi.org/10.1016/j.ijid.2010.03.022

Quan, T.M., Thao, T.T.N., Duy, N.M., Nhat, T.M., Clapham, H., 2020. Estimates of the global burden of Japanese encephalitis and the impact of vaccination from 2000-2015. eLife 9, e51027. https://doi.org/10.7554/eLife.51027

Sohn, Y.M., 2000. Japanese Encephalitis Immunization in South Korea: Past, Present, and Future. Emerging Infectious Diseases 6, 8.

WHO, 2016. Global Immunization News. March, 2016. URL: https://www.who.int/immunization/GIN_March_2016.pdf (accessed 4.6.21)

WHO, 2018. Japanese encephalitis vaccination campaign achievement [WWW Document]. URL: https://www.who.int/myanmar/our-work/japanese-encephalitis-vaccination-campaign-achievement (accessed 4.6.21).

WHO, 2020. WHO World Health Organization: Immunization, Vaccines And Biologicals. Vaccine preventable diseases Vaccines monitoring system 2020 Global Summary Reference Time Series: JAPENC [WWW Document]. URL: https://apps.who.int/immunization_monitoring/globalsummary/timeseries/tscoveragejapenc.html (accessed 1.5.21).

**Table S3. Mean annual number of JE cases (C) and deaths (D) per country from 2010 to 2019 under the vaccination and no vaccination scenarios.** Numbers are presented for three different estimates of the at-risk population size - baseline, low, and high. See Methods for description of how these population sizes were estimated.

| | | Vaccination | | | No Vaccination | | |
|---|---|---|---|---|---|---|---|
| Country | | Baseline (95% CI) | Low (95% CI) | High (95% CI) | Baseline (95% CI) | Low (95% CI) | High (95% CI) |
| Bangladesh | C | 9476 (2958 - 31275) | 9435 (2952 - 30980) | 11581 (3629 - 38044) | 9476 (2964 - 31047) | 9434 (2956 - 30894) | 11580 (3634 - 37867) |
| | D | 3447 (372 - 13209) | 3431 (368 - 13173) | 4211 (455 - 16230) | 3446 (377 - 13218) | 3431 (370 - 13217) | 4210 (458 - 16199) |
| Bhutan | C | 2 (0 - 7) | 1 (0 - 3) | 7 (1 - 22) | 2 (0 - 7) | 1 (0 - 3) | 7 (0 - 22) |
| | D | 1 (0 - 3) | 0 (0 - 2) | 2 (0 - 10) | 1 (0 - 3) | 0 (0 - 2) | 2 (0 - 10) |
| Brunei | C | 4 (0 - 14) | 2 (0 - 9) | 7 (1 - 24) | 4 (0 - 14) | 3 (0 - 9) | 7 (1 - 24) |
| | D | 1 (0 - 7) | 1 (0 - 4) | 3 (0 - 11) | 1 (0 - 7) | 1 (0 - 5) | 3 (0 - 11) |
| Cambodia | C | 431 (76 - 1458) | 408 (71 - 1383) | 638 (113 - 2141) | 596 (185 - 1996) | 565 (173 - 1876) | 882 (275 - 2932) |
| | D | 157 (11 - 641) | 148 (10 - 607) | 232 (17 - 950) | 217 (22 - 816) | 206 (21 - 775) | 321 (33 - 1210) |
| China | C | 10113 (1415 - 35180) | 10071 (1405 - 35014) | 11231 (1568 - 39057) | 24953 (7783 - 82755) | 24849 (7768 - 82222) | 27705 (8642 - 91831) |
| | D | 3642 (237 - 14890) | 3628 (237 - 14867) | 4045 (265 - 16587) | 9071 (981 - 34878) | 9034 (989 - 34631) | 10072 (1087 - 38914) |
| India | C | 20342 (6277 - 65953) | 12282 (3800 - 39895) | 30563 (9463 - 98755) | 23980 (7428 - 80043) | 14478 (4491 - 48374) | 36029 (11146 - 120784) |
| | D | 7399 (791 - 28080) | 4467 (484 - 16950) | 11118 (1202 - 42214) | 8725 (938 - 34227) | 5268 (566 - 20611) | 13108 (1411 - 51169) |
| Indonesia | C | 4815 (1506 - 15935) | 3528 (1099 - 11672) | 8679 (2719 - 28739) | 4826 (1508 - 15967) | 3538 (1104 - 11655) | 8698 (2709 - 28856) |
| | D | 1752 (188 - 6780) | 1283 (140 - 4952) | 3157 (345 - 12091) | 1756 (191 - 6773) | 1287 (140 - 4957) | 3164 (343 - 12149) |
| Japan | C | 353 (94 - 1177) | 352 (94 - 1171) | 376 (101 - 1257) | 506 (130 - 1692) | 504 (132 - 1666) | 538 (140 - 1792) |
| | D | 129 (12 - 494) | 128 (12 - 496) | 137 (13 - 529) | 184 (18 - 720) | 183 (18 - 711) | 196 (19 - 760) |
| Laos | C | 102 (29 - 322) | 94 (26 - 293) | 121 (34 - 380) | 105 (30 - 330) | 97 (27 - 307) | 125 (35 - 392) |
| | D | 37 (3 - 142) | 34 (3 - 129) | 44 (4 - 163) | 38 (3 - 144) | 35 (3 - 131) | 45 (4 - 170) |
| Malaysia | C | 106 (10 - 421) | 69 (6 - 273) | 162 (17 - 650) | 541 (170 - 1777) | 353 (108 - 1167) | 828 (257 - 2705) |
| | D | 39 (1 - 196) | 26 (1 - 128) | 60 (2 - 296) | 197 (20 - 759) | 128 (13 - 494) | 301 (31 - 1151) |
| Myanmar | C | 1028 (155 - 3458) | 973 (148 - 3247) | 1376 (205 - 4613) | 1268 (383 - 4264) | 1200 (361 - 4045) | 1697 (510 - 5766) |
| | D | 373 (26 - 1483) | 353 (24 - 1401) | 499 (34 - 1978) | 460 (47 - 1697) | 435 (43 - 1599) | 615 (63 - 2275) |
| Nepal | C | 570 (146 - 1819) | 344 (89 - 1100) | 654 (171 - 2086) | 940 (294 - 3105) | 567 (176 - 1862) | 1079 (338 - 3575) |
| | D | 207 (19 - 782) | 125 (11 - 475) | 238 (22 - 886) | 342 (36 - 1292) | 206 (22 - 777) | 393 (42 - 1481) |
| North Korea | C | 257 (74 - 820) | 257 (72 - 820) | 288 (82 - 923) | 361 (107 - 1219) | 361 (106 - 1207) | 404 (121 - 1352) |
| | D | 94 (9 - 358) | 94 (9 - 363) | 105 (10 - 399) | 132 (13 - 508) | 131 (14 - 517) | 147 (15 - 565) |
| Pakistan | C | 1179 (301 - 3877) | 483 (122 - 1603) | 4341 (1107 - 14339) | 1179 (301 - 3881) | 483 (123 - 1587) | 4342 (1111 - 14300) |
| | D | 430 (40 - 1686) | 176 (16 - 698) | 1581 (153 - 6255) | 429 (40 - 1677) | 176 (16 - 697) | 1581 (152 - 6246) |
| Papua New | C | 33 (6 - 115) | 1 (0 - 4) | 43 (9 - 144) | 33 (6 - 112) | 1 (0 - 4) | 43 (9 - 145) |
| Guinea | D | 12 (0 - 48) | 0 (0 - 2) | 15 (1 - 62) | 12 (0 - 48) | 0 (0 - 2) | 16 (1 - 61) |
| Philippines | C | 2080 (638 - 6607) | 2010 (617 - 6372) | 3263 (1008 - 10383) | 2080 (636 - 6566) | 2010 (620 - 6379) | 3263 (1008 - 10405) |
| | D | 758 (79 - 2959) | 734 (76 - 2851) | 1190 (123 - 4612) | 759 (78 - 2975) | 733 (76 - 2837) | 1190 (126 - 4603) |
| Singapore | C | 26 (6 - 82) | 26 (6 - 82) | 61 (15 - 193) | 26 (6 - 83) | 26 (6 - 84) | 61 (16 - 189) |
| | D | 9 (0 - 37) | 9 (0 - 37) | 22 (1 - 89) | 9 (0 - 38) | 9 (0 - 37) | 22 (1 - 87) |
| South Korea | C | 114 (31 - 368) | 107 (28 - 345) | 141 (38 - 451) | 505 (155 - 1654) | 474 (145 - 1565) | 622 (192 - 2039) |

| | | | | | | | | | | | | | |
|---|---|---|---|---|---|---|---|---|---|---|---|---|---|
| | D | 41 | (3 - 157) | 39 | (3 - 148) | 51 | (4 - 196) | 184 | (19 - 689) | 173 | (17 - 654) | 226 | (23 - 861) |
| Sri Lanka | C | 14 | (2 - 44) | 5 | (0 - 17) | 76 | (21 - 242) | 52 | (14 - 172) | 20 | (5 - 67) | 291 | (90 - 964) |
| | D | 5 | (0 - 20) | 2 | (0 - 8) | 28 | (2 - 104) | 19 | (1 - 74) | 7 | (0 - 29) | 106 | (11 - 414) |
| Taiwan | C | 6 | (0 - 20) | 6 | (0 - 20) | 14 | (2 - 44) | 68 | (19 - 223) | 68 | (20 - 221) | 157 | (47 - 511) |
| | D | 2 | (0 - 9) | 2 | (0 - 9) | 5 | (0 - 20) | 25 | (2 - 95) | 25 | (2 - 96) | 57 | (5 - 220) |
| Thailand | C | 280 | (69 - 932) | 263 | (65 - 879) | 548 | (136 - 1823) | 915 | (287 - 3022) | 859 | (269 - 2844) | 1792 | (562 - 5896) |
| | D | 102 | (8 - 425) | 96 | (8 - 399) | 200 | (18 - 822) | 333 | (35 - 1289) | 313 | (33 - 1204) | 651 | (71 - 2503) |
| Timor-Leste | C | 1 | (0 - 3) | 0 | (0 - 2) | 10 | (1 - 34) | 1 | (0 - 3) | 0 | (0 - 2) | 10 | (1 - 34) |
| | D | 0 | (0 - 2) | 0 | (0 - 1) | 4 | (0 - 16) | 0 | (0 - 2) | 0 | (0 - 1) | 4 | (0 - 15) |
| Vietnam | C | 1456 | (345 - 4739) | 1444 | (342 - 4714) | 1882 | (450 - 6113) | 3332 | (1046 - 10997) | 3302 | (1033 - 10915) | 4308 | (1349 - 14197) |
| | D | 530 | (48 - 2049) | 526 | (47 - 2044) | 685 | (62 - 2643) | 1212 | (131 - 4647) | 1201 | (132 - 4640) | 1566 | (167 - 6060) |

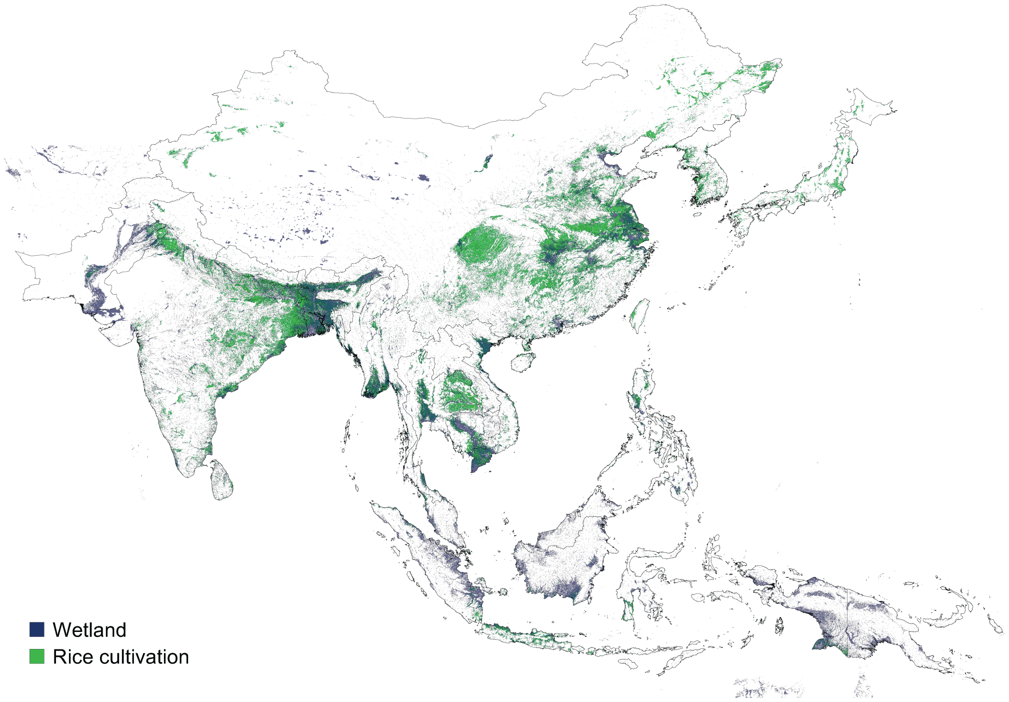


**SI Fig 1. Spatial extent of rice cultivation (green) and wetlands (blue) in Asia.** Rice cultivation map is derived from [9]. Extent of wetlands (including seasonal wetlands) based on wetlands types in tropical and sub-tropical Asia from [10].

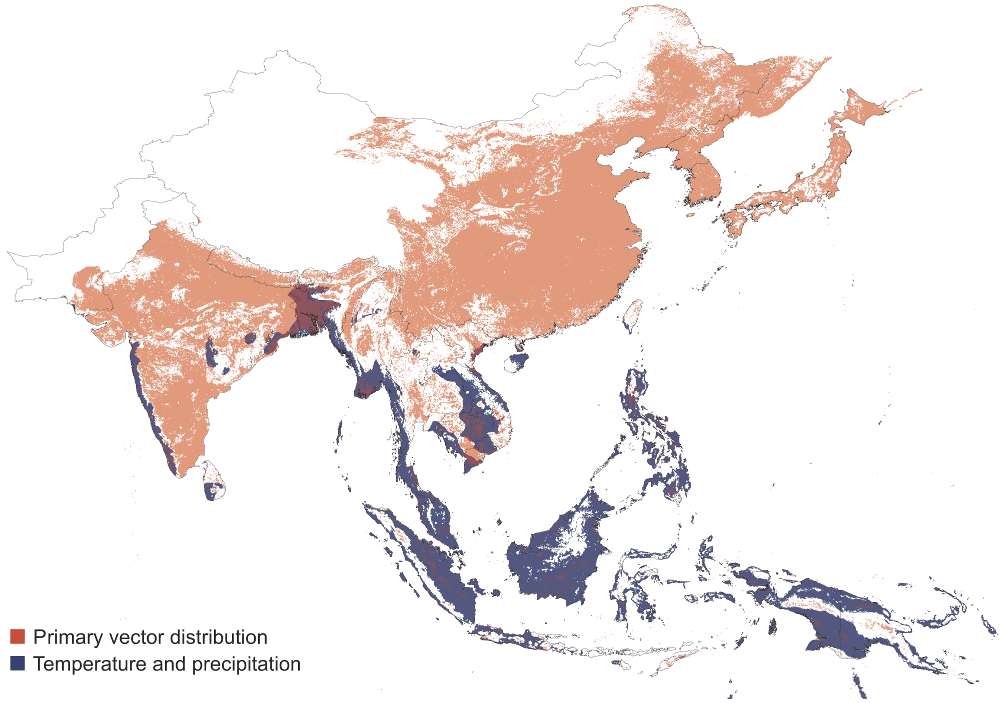


**SI Fig 2. Map of likely suitable habitat for JEV vector species.** Orange represents areas where the modeled probability of *Culex tritaeniorhynchus* occurrence is $\geq 0.25$ according to [12]. Blue represents regions with an annual mean temp of $\geq 20°C$ and $\geq 1500$mm of annual precipitation. Darker orange areas represent regions where these two distributions overlap.

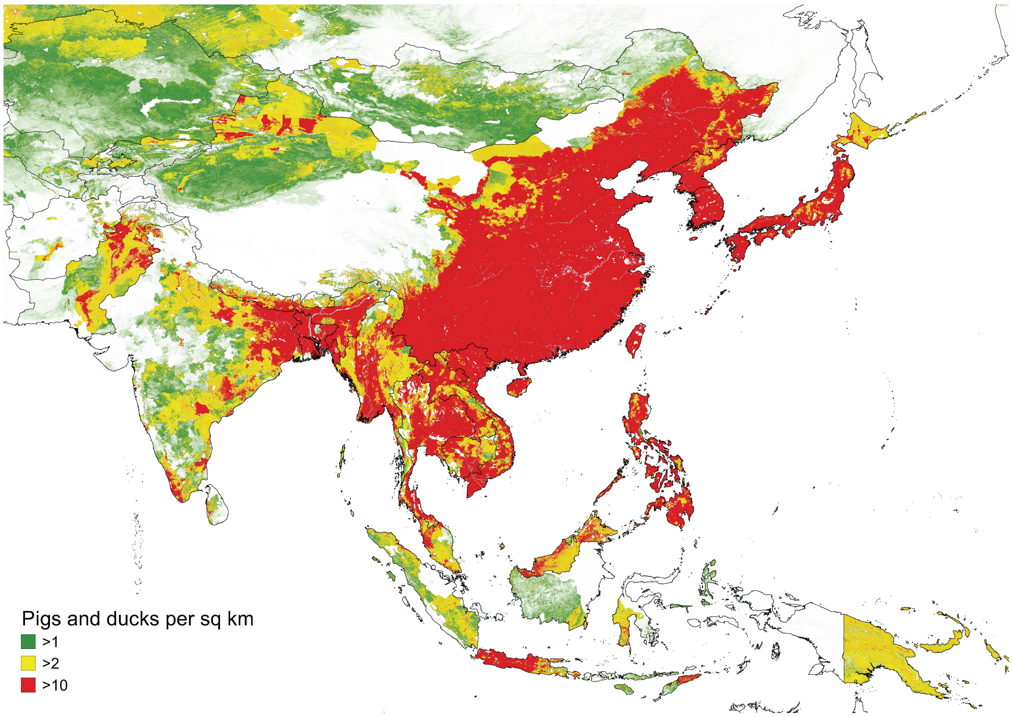


**SI Fig 3. Combined population densities per kilometer of domestic pigs and ducks.** Map shows the three different thresholds used in estimating the size of the at-risk population in JE-endemic areas. The default threshold used for sustained JEV transmission was $\geq 2$ per square km. Derived from [17].

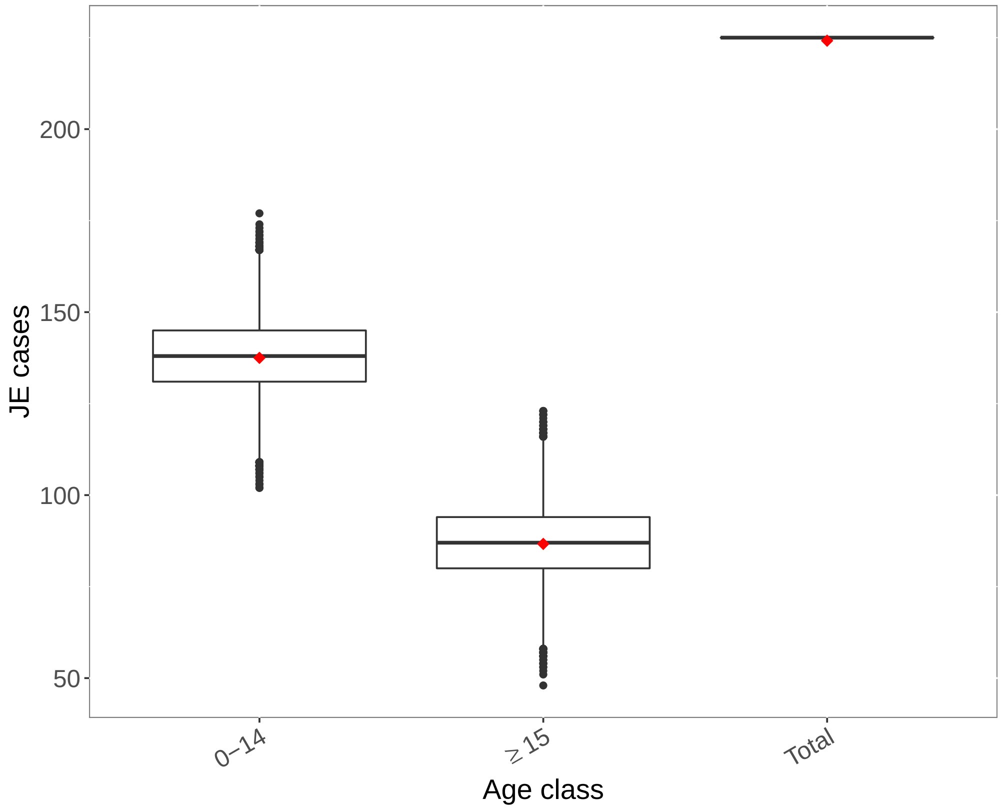


**SI Fig 4. Observed versus predicted number of JE cases per age class in Bangladesh based on dataset from [15].** Boxplots represent predicted number of cases per age class based on draws from the joint posterior distribution of FOI and vaccination coverage (if included) estimates. Red diamonds represent the observed number of cases.

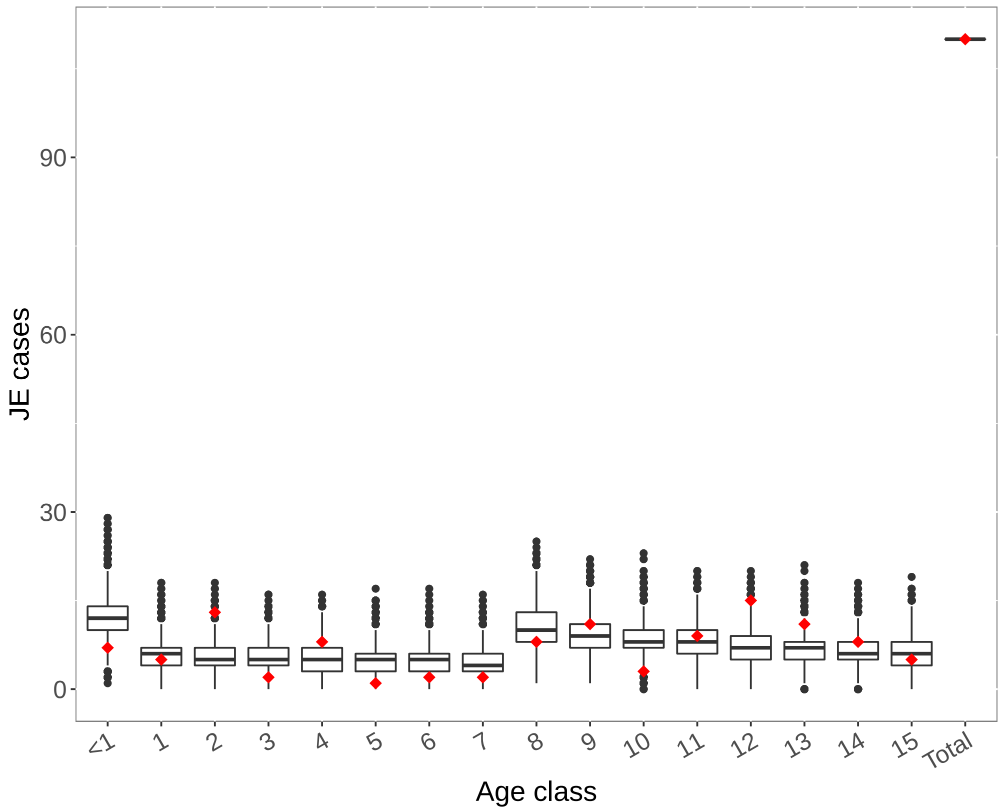


**SI Fig 5.** Observed versus predicted number of JE cases per age class in Cambodia based on dataset from [41]. Boxplots represent predicted number of cases per age class based on draws from the joint posterior distribution of FOI and vaccination coverage (if included) estimates. Red diamonds represent the observed number of cases.

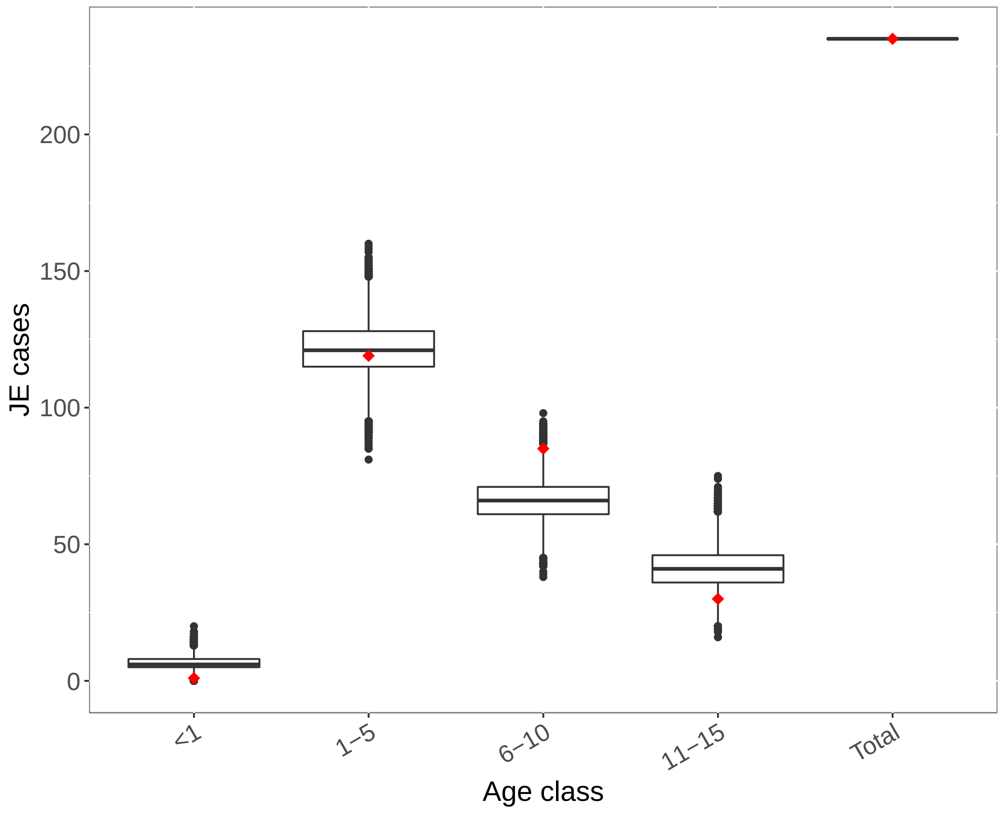


**SI Fig 6.** Observed versus predicted number of JE cases per age class in Cambodia based on dataset from [42]. Boxplots represent predicted number of cases per age class based on draws from the joint posterior distribution of FOI and vaccination coverage (if included) estimates. Red diamonds represent the observed number of cases.

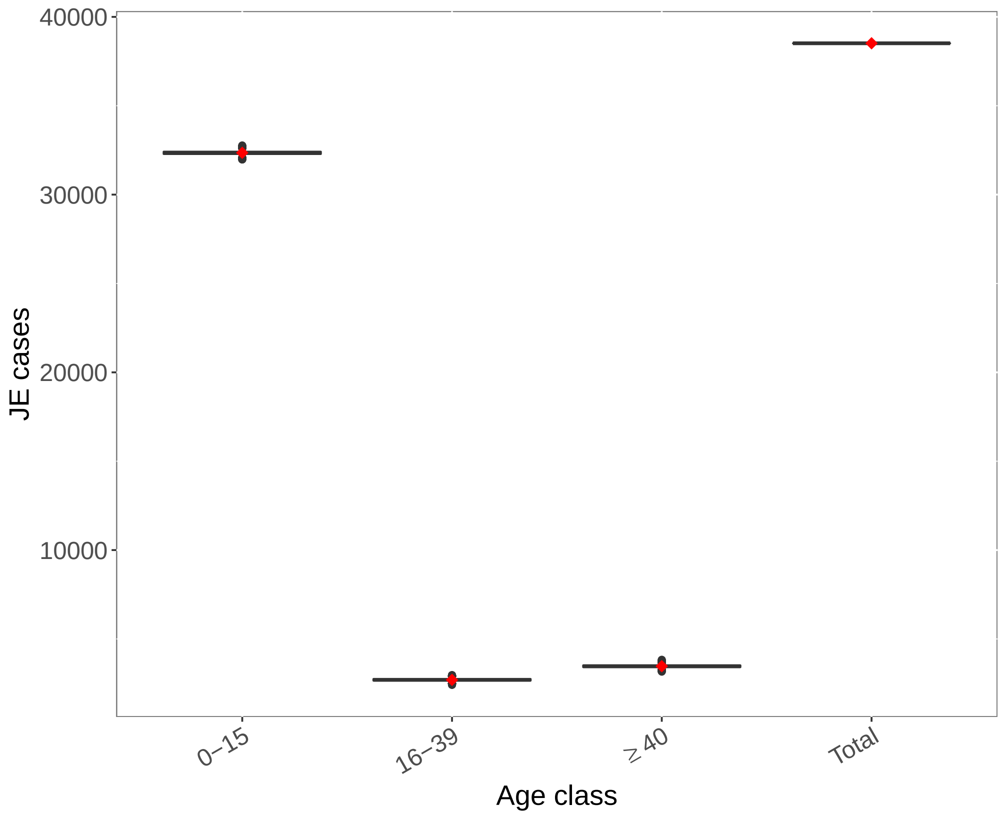


**SI Fig 7.** Observed versus predicted number of JE cases per age class in China based on dataset from [43]. Boxplots represent predicted number of cases per age class based on draws from the joint posterior distribution of FOI and vaccination coverage (if included) estimates. Red diamonds represent the observed number of cases.

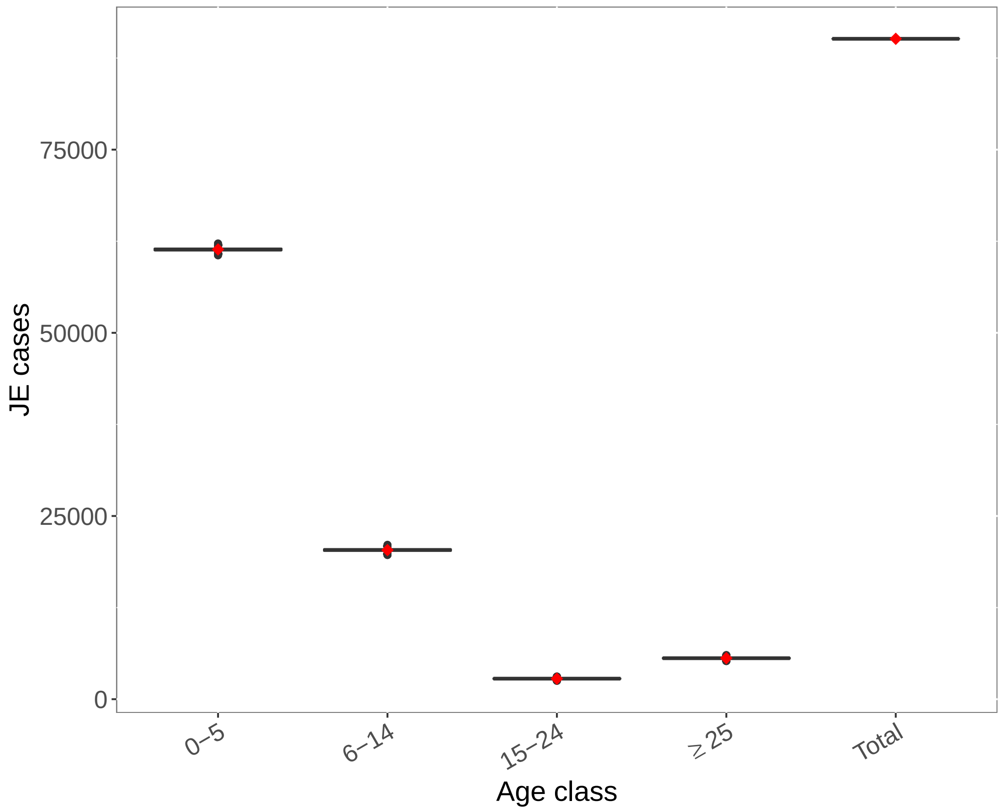


**SI Fig 8.** Observed versus predicted number of JE cases per age class in China based on dataset from [44]. Boxplots represent predicted number of cases per age class based on draws from the joint posterior distribution of FOI and vaccination coverage (if included) estimates. Red diamonds represent the observed number of cases.

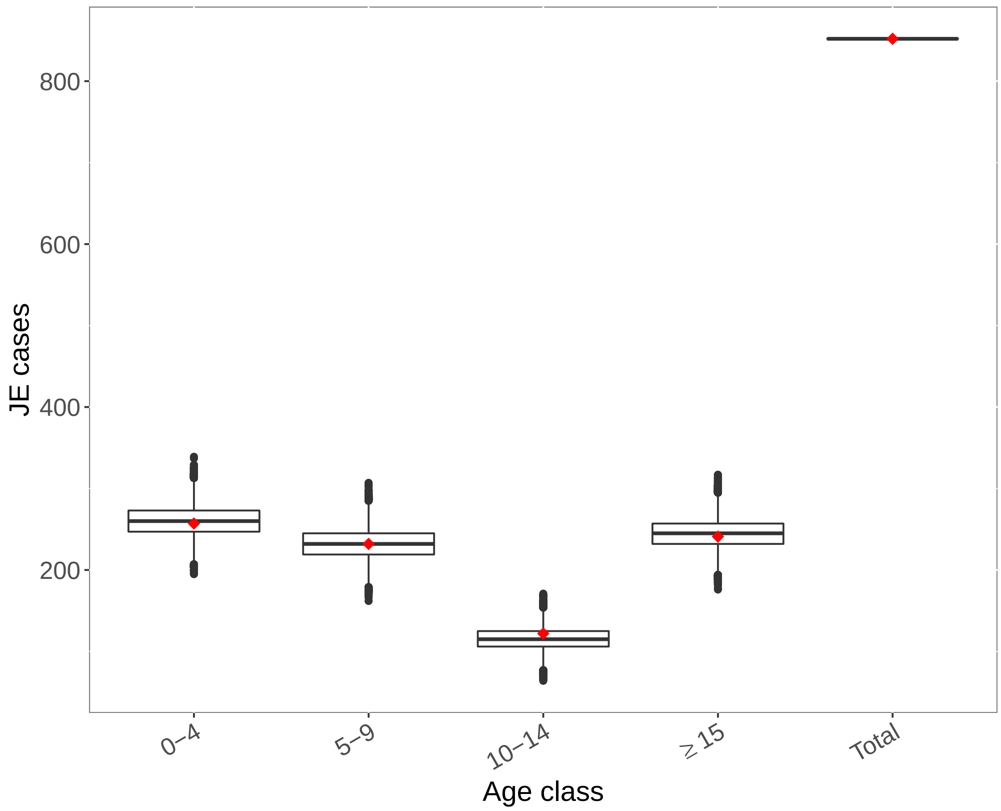


**SI Fig 9.** Observed versus predicted number of JE cases per age class in India based on dataset from [45]. Boxplots represent predicted number of cases per age class based on draws from the joint posterior distribution of FOI and vaccination coverage (if included) estimates. Red diamonds represent the observed number of cases.

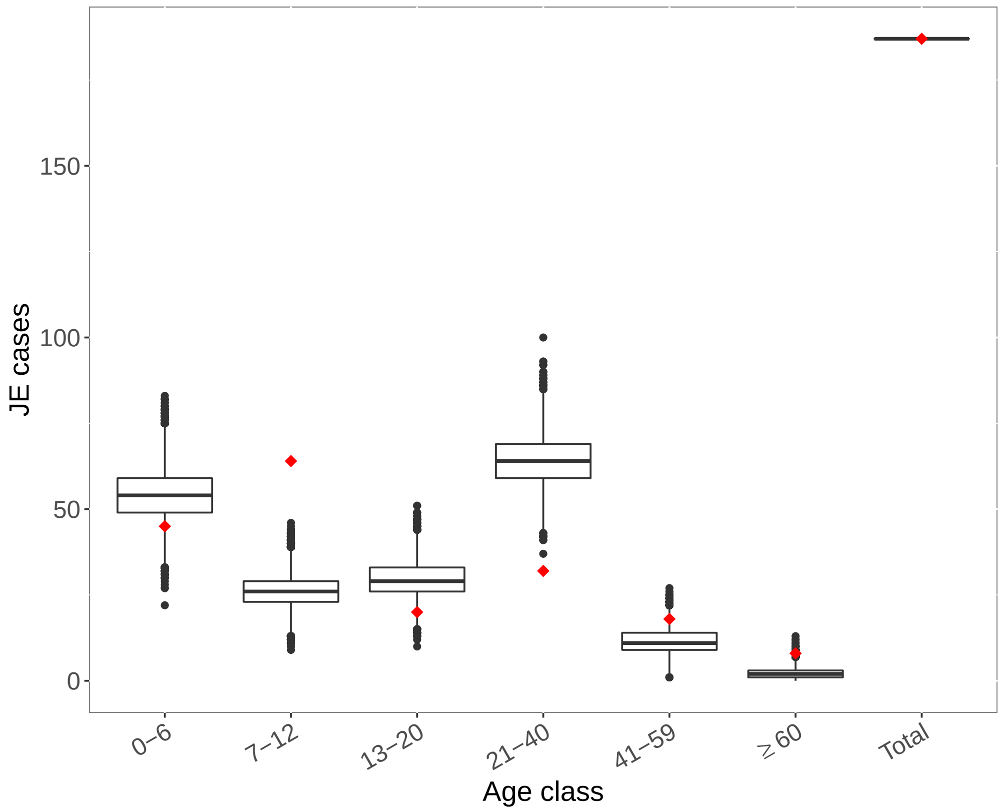


**SI Fig 10.** Observed versus predicted number of JE cases per age class in India based on dataset from [46]. Boxplots represent predicted number of cases per age class based on draws from the joint posterior distribution of FOI and vaccination coverage (if included) estimates. Red diamonds represent the observed number of cases.

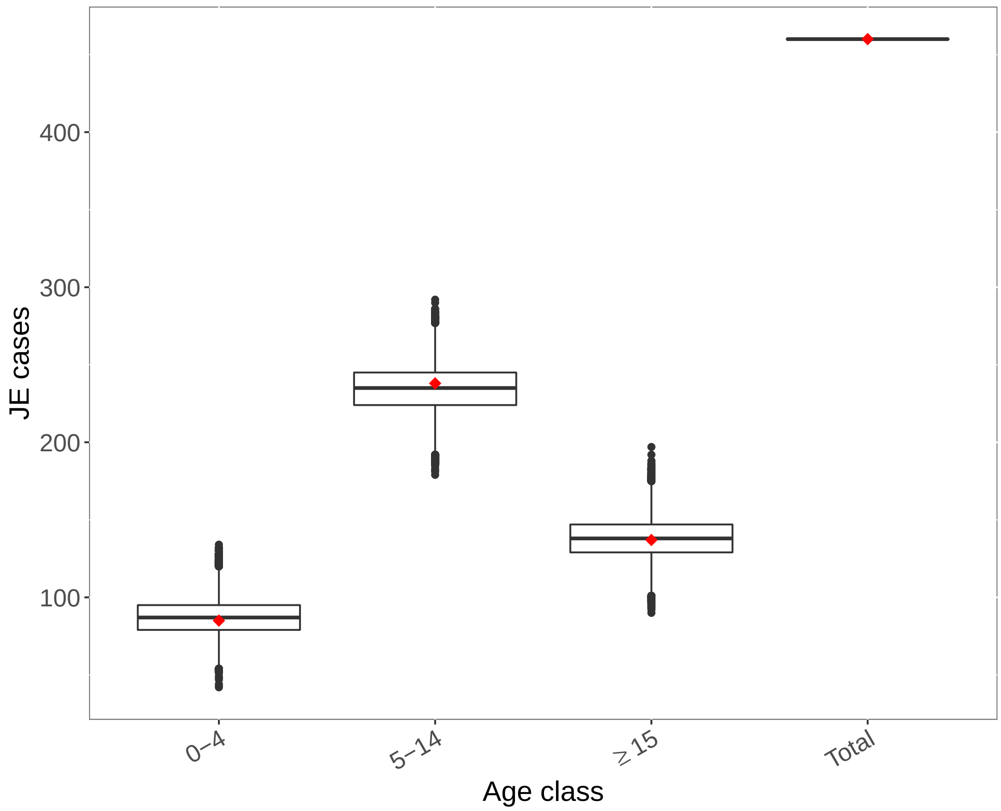


**SI Fig 11.** Observed versus predicted number of JE cases per age class in India based on dataset from [47]. Boxplots represent predicted number of cases per age class based on draws from the joint posterior distribution of FOI and vaccination coverage (if included) estimates. Red diamonds represent the observed number of cases.

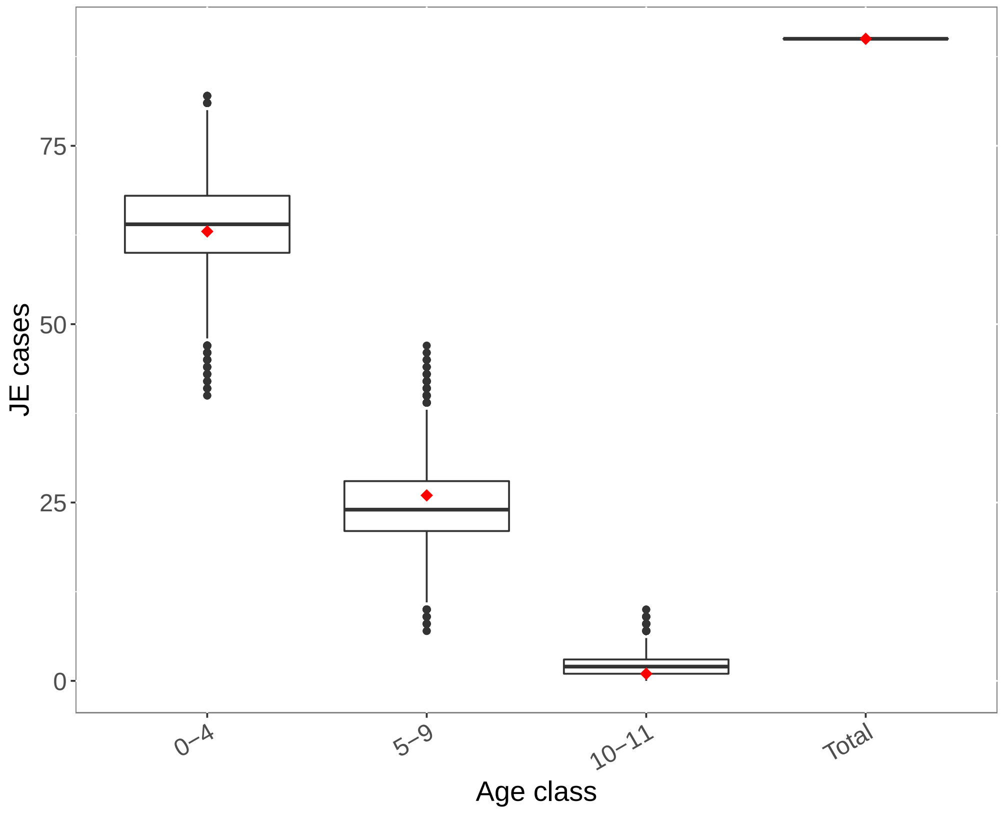


**SI Fig 12.** Observed versus predicted number of JE cases per age class in Indonesia based on dataset from [48]. Boxplots represent predicted number of cases per age class based on draws from the joint posterior distribution of FOI and vaccination coverage (if included) estimates. Red diamonds represent the observed number of cases.

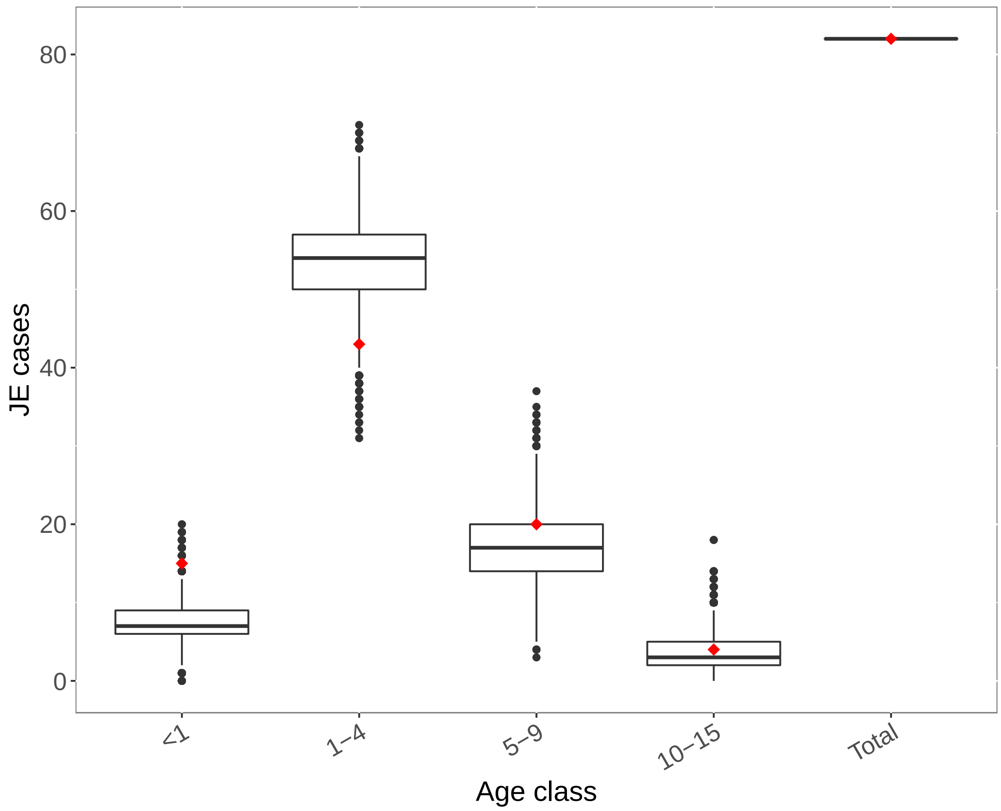


**SI Fig 13.** Observed versus predicted number of JE cases per age class in Indonesia based on dataset from [16]. Boxplots represent predicted number of cases per age class based on draws from the joint posterior distribution of FOI and vaccination coverage (if included) estimates. Red diamonds represent the observed number of cases.

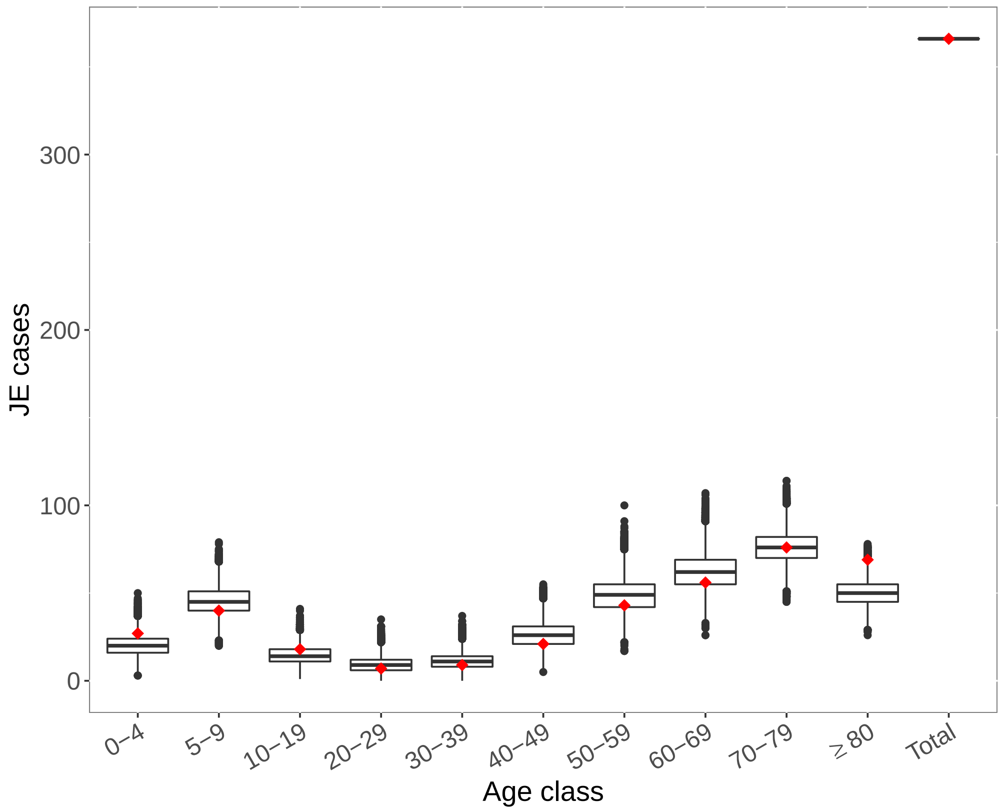


**SI Fig 14.** Observed versus predicted number of JE cases per age class in Japan based on dataset from [49]. Boxplots represent predicted number of cases per age class based on draws from the joint posterior distribution of FOI and vaccination coverage (if included) estimates. Red diamonds represent the observed number of cases.

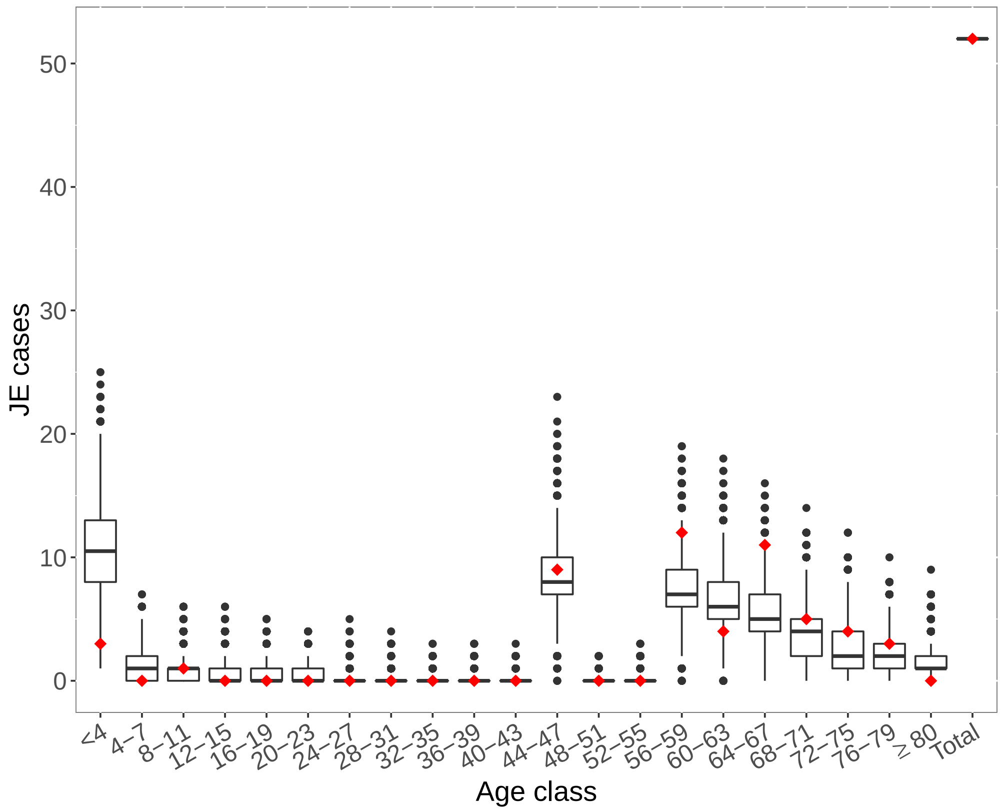


**SI Fig 15.** Observed versus predicted number of JE cases per age class in Laos based on dataset from [50]. Boxplots represent predicted number of cases per age class based on draws from the joint posterior distribution of FOI and vaccination coverage (if included) estimates. Red diamonds represent the observed number of cases.

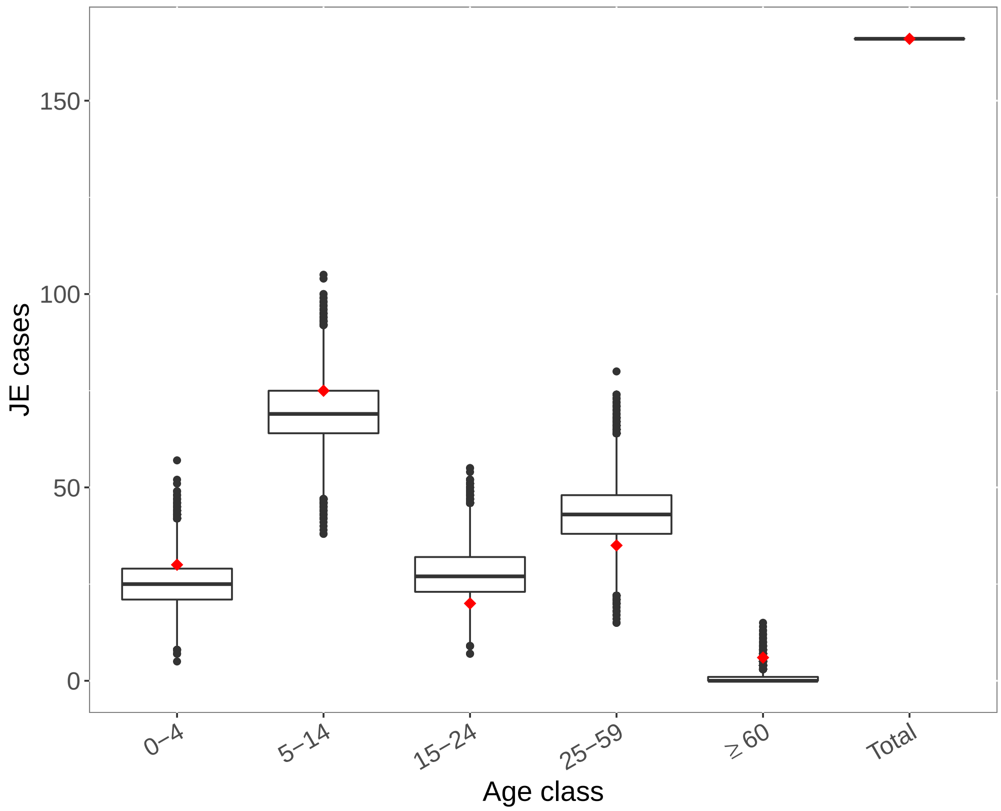


**SI Fig 16.** Observed versus predicted number of JE cases per age class in Malaysia based on dataset from [51]. Boxplots represent predicted number of cases per age class based on draws from the joint posterior distribution of FOI and vaccination coverage (if included) estimates. Red diamonds represent the observed number of cases.

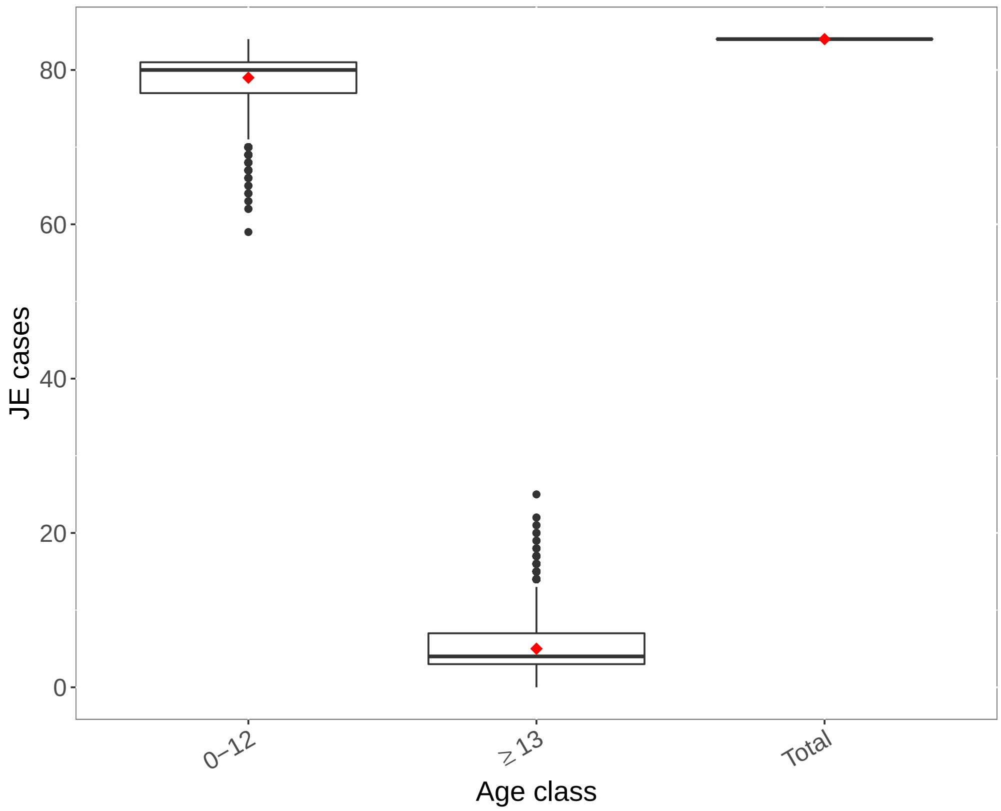


**SI Fig 17.** Observed versus predicted number of JE cases per age class in Malaysia based on dataset from [19]. Boxplots represent predicted number of cases per age class based on draws from the joint posterior distribution of FOI and vaccination coverage (if included) estimates. Red diamonds represent the observed number of cases.

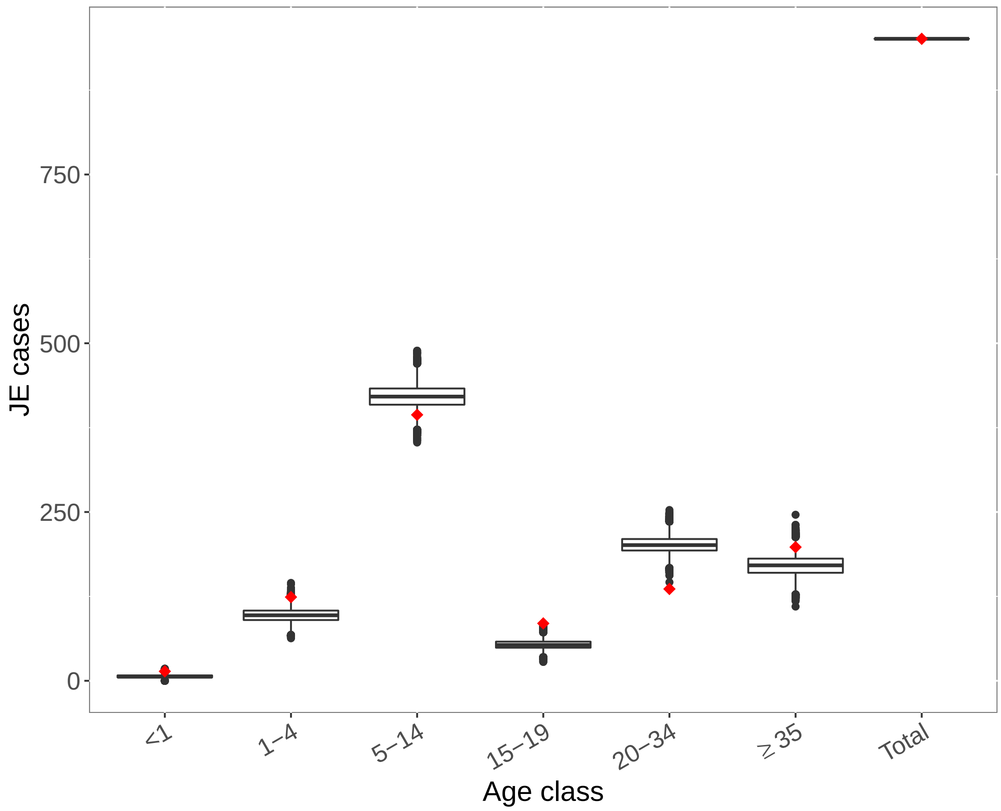


**SI Fig 18.** Observed versus predicted number of JE cases per age class in Nepal based on dataset from [52]. Boxplots represent predicted number of cases per age class based on draws from the joint posterior distribution of FOI and vaccination coverage (if included) estimates. Red diamonds represent the observed number of cases.

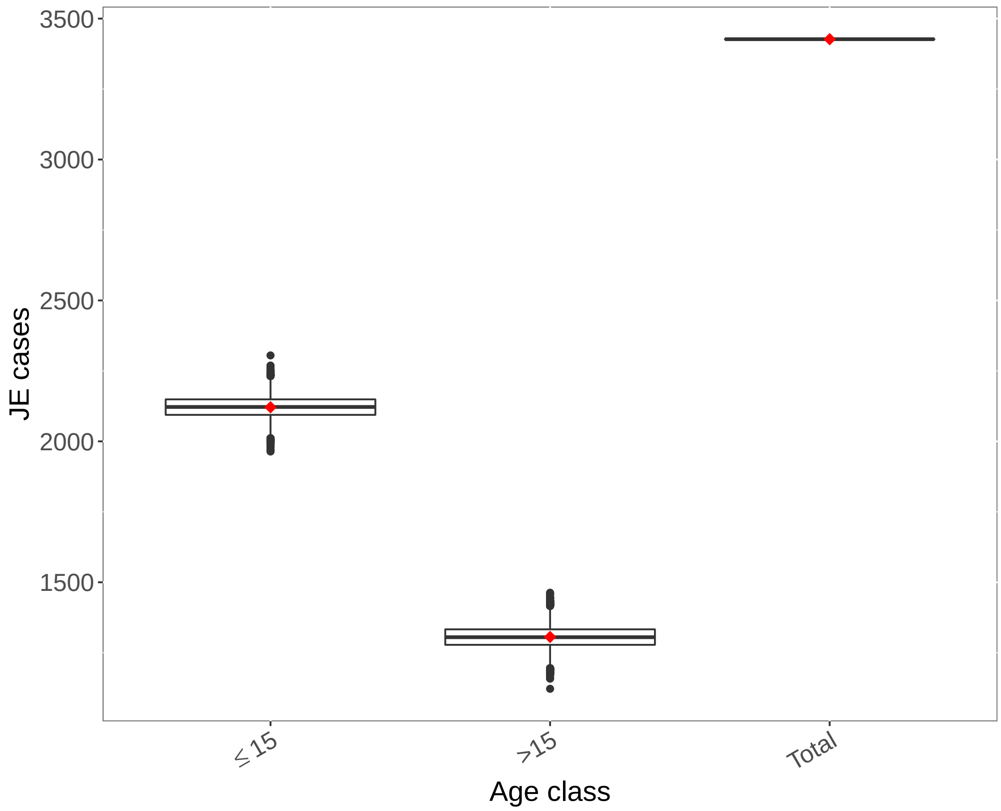


**SI Fig 19.** Observed versus predicted number of JE cases per age class in Nepal based on dataset from [53]. Boxplots represent predicted number of cases per age class based on draws from the joint posterior distribution of FOI and vaccination coverage (if included) estimates. Red diamonds represent the observed number of cases.

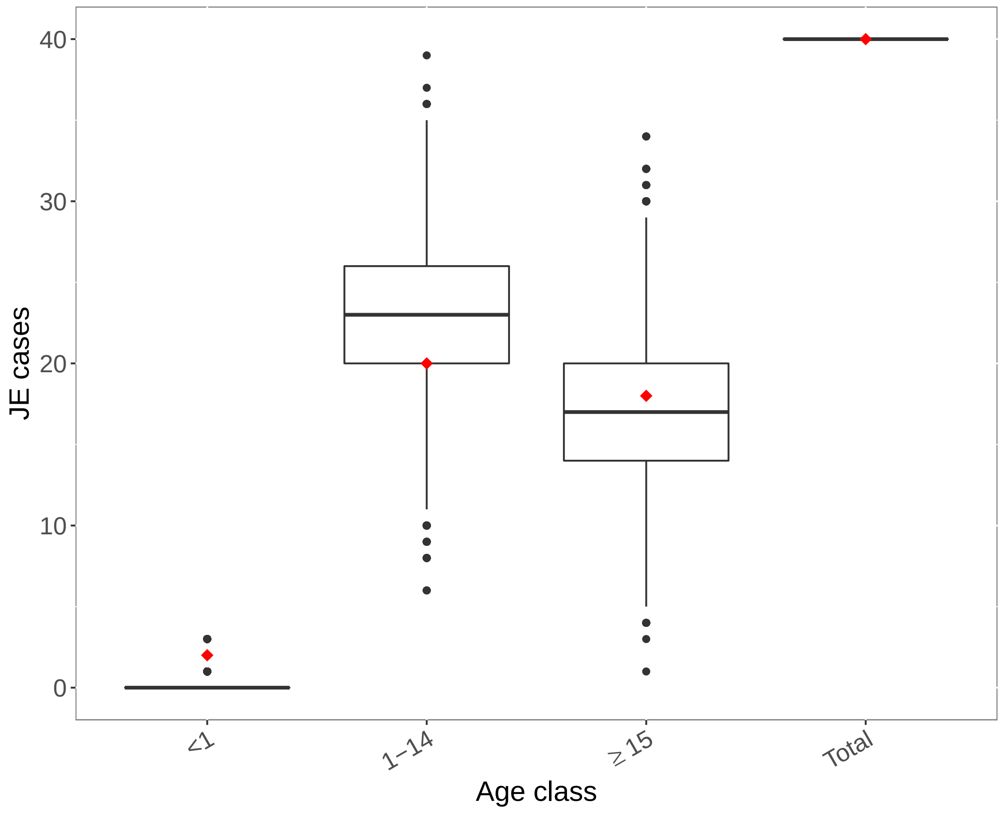


**SI Fig 20.** Observed versus predicted number of JE cases per age class in Nepal based on dataset from [54]. Boxplots represent predicted number of cases per age class based on draws from the joint posterior distribution of FOI and vaccination coverage (if included) estimates. Red diamonds represent the observed number of cases.

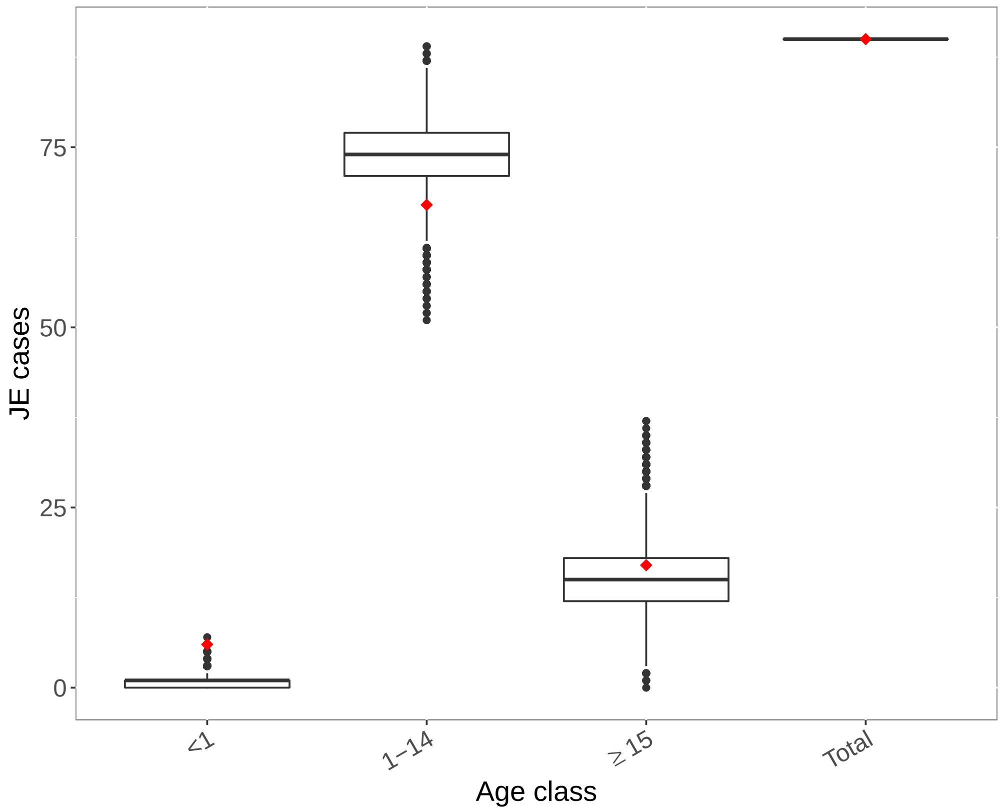


**SI Fig 21.** Observed versus predicted number of JE cases per age class in Nepal based on dataset from [55]. Boxplots represent predicted number of cases per age class based on draws from the joint posterior distribution of FOI and vaccination coverage (if included) estimates. Red diamonds represent the observed number of cases.

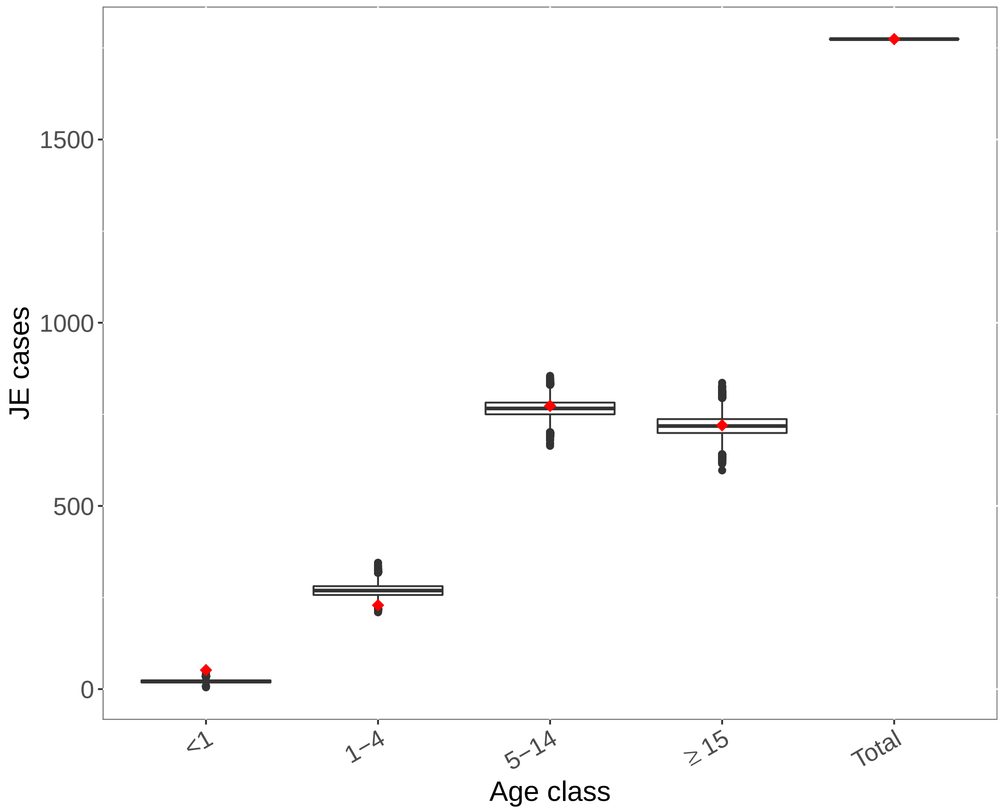


**SI Fig 22.** Observed versus predicted number of JE cases per age class in Nepal based on dataset from [18]. Boxplots represent predicted number of cases per age class based on draws from the joint posterior distribution of FOI and vaccination coverage (if included) estimates. Red diamonds represent the observed number of cases.

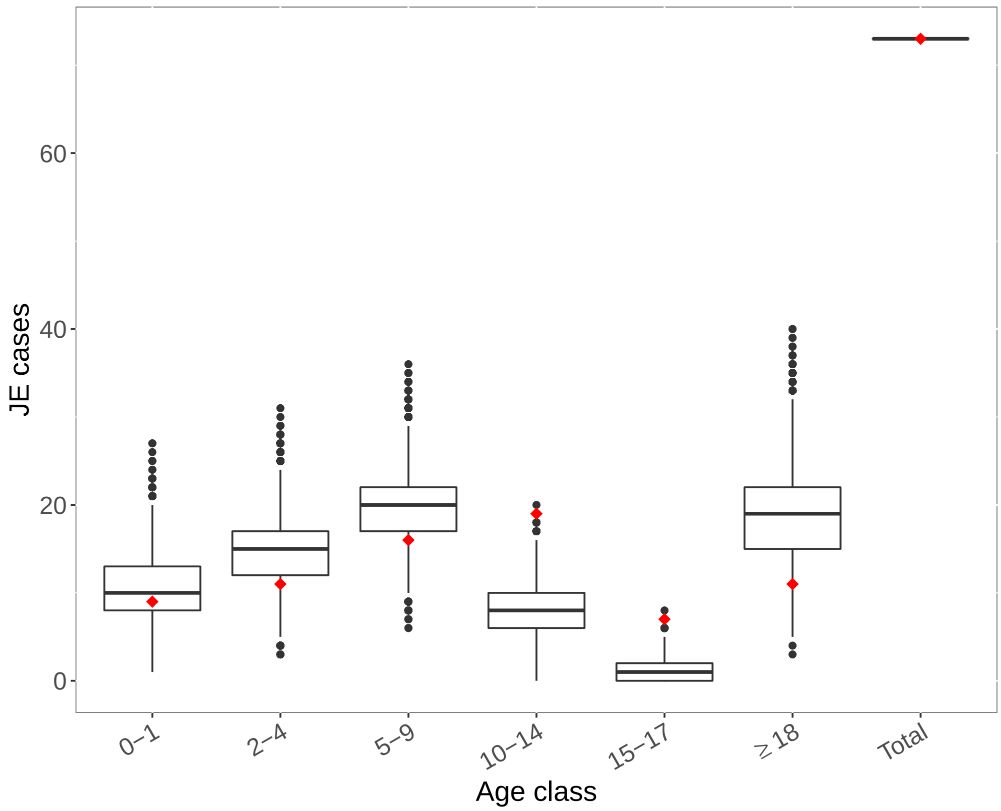


**SI Fig 23.** Observed versus predicted number of JE cases per age class in the Philippines based on dataset from [56]. Boxplots represent predicted number of cases per age class based on draws from the joint posterior distribution of FOI and vaccination coverage (if included) estimates. Red diamonds represent the observed number of cases.

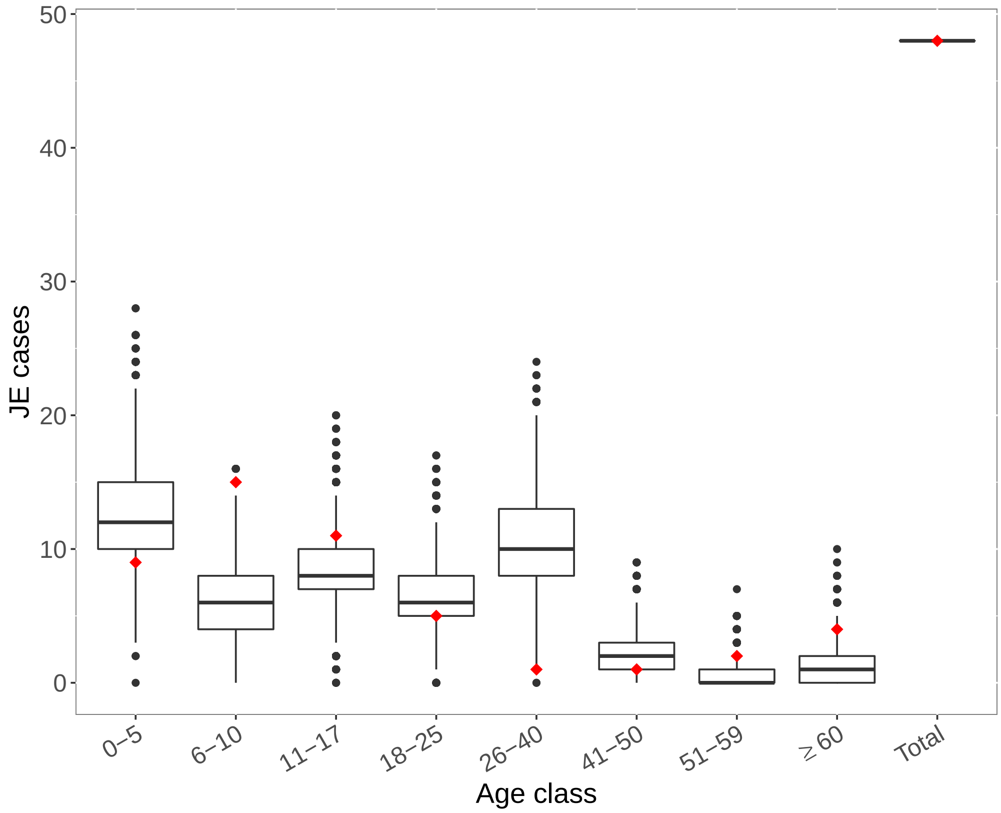


**SI Fig 24.** Observed versus predicted number of JE cases per age class in the Philippines based on dataset from [57]. Boxplots represent predicted number of cases per age class based on draws from the joint posterior distribution of FOI and vaccination coverage (if included) estimates. Red diamonds represent the observed number of cases.

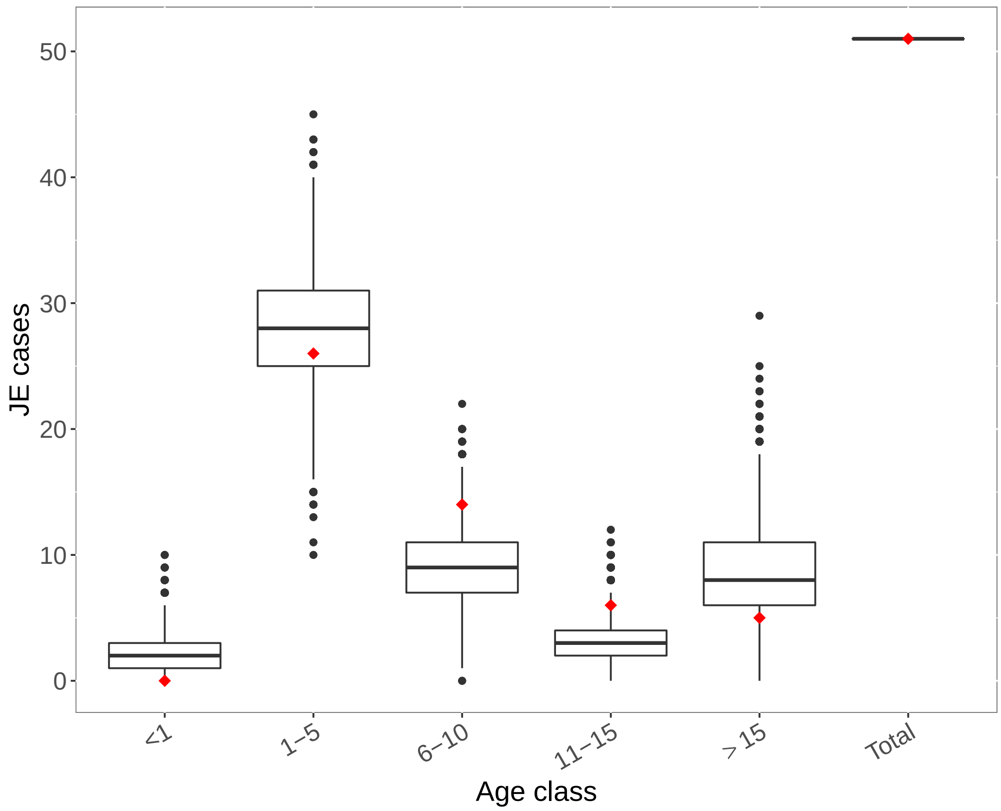


**SI Fig 25.** Observed versus predicted number of JE cases per age class in the Philippines based on dataset from [58]. Boxplots represent predicted number of cases per age class based on draws from the joint posterior distribution of FOI and vaccination coverage (if included) estimates. Red diamonds represent the observed number of cases.

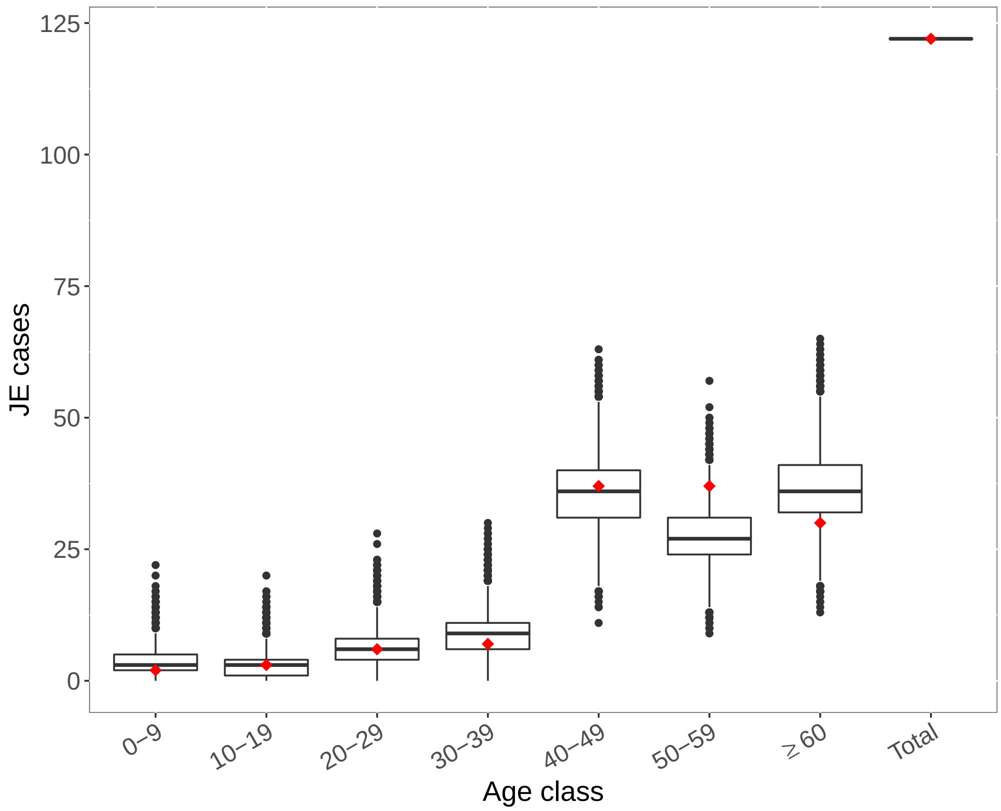


**SI Fig 26.** Observed versus predicted number of JE cases per age class in South Korea based on dataset from [59]. Boxplots represent predicted number of cases per age class based on draws from the joint posterior distribution of FOI and vaccination coverage (if included) estimates. Red diamonds represent the observed number of cases.

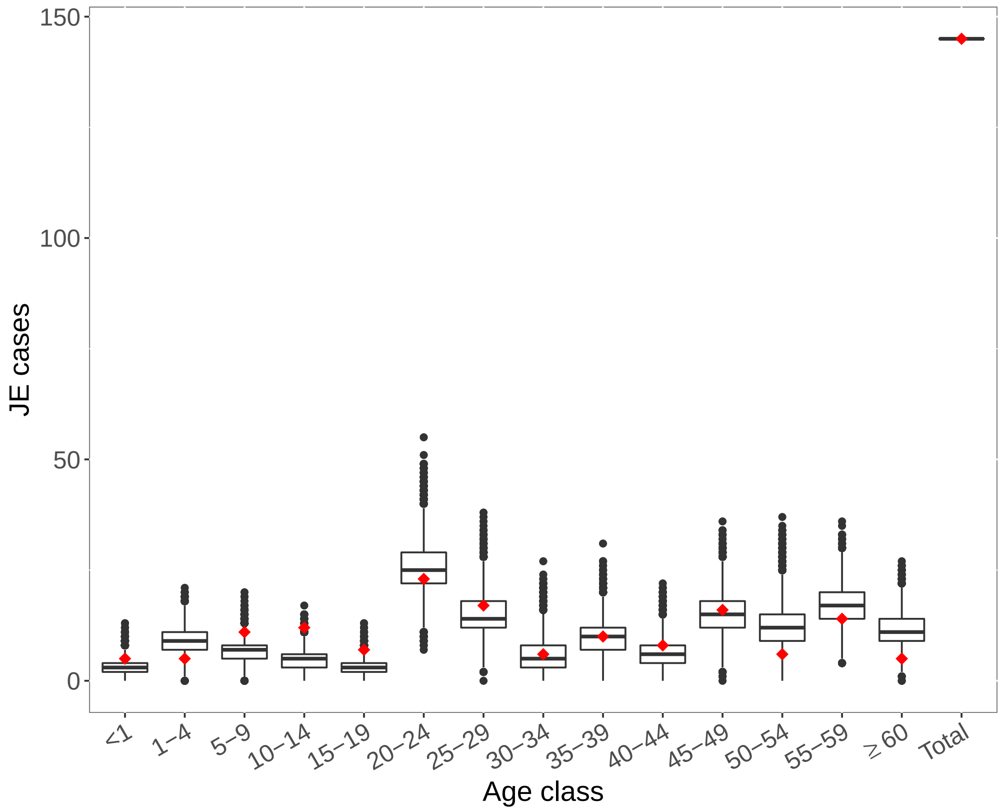


**SI Fig 27.** Observed versus predicted number of JE cases per age class in Sri Lanka based on dataset from [60]. Boxplots represent predicted number of cases per age class based on draws from the joint posterior distribution of FOI and vaccination coverage (if included) estimates. Red diamonds represent the observed number of cases.

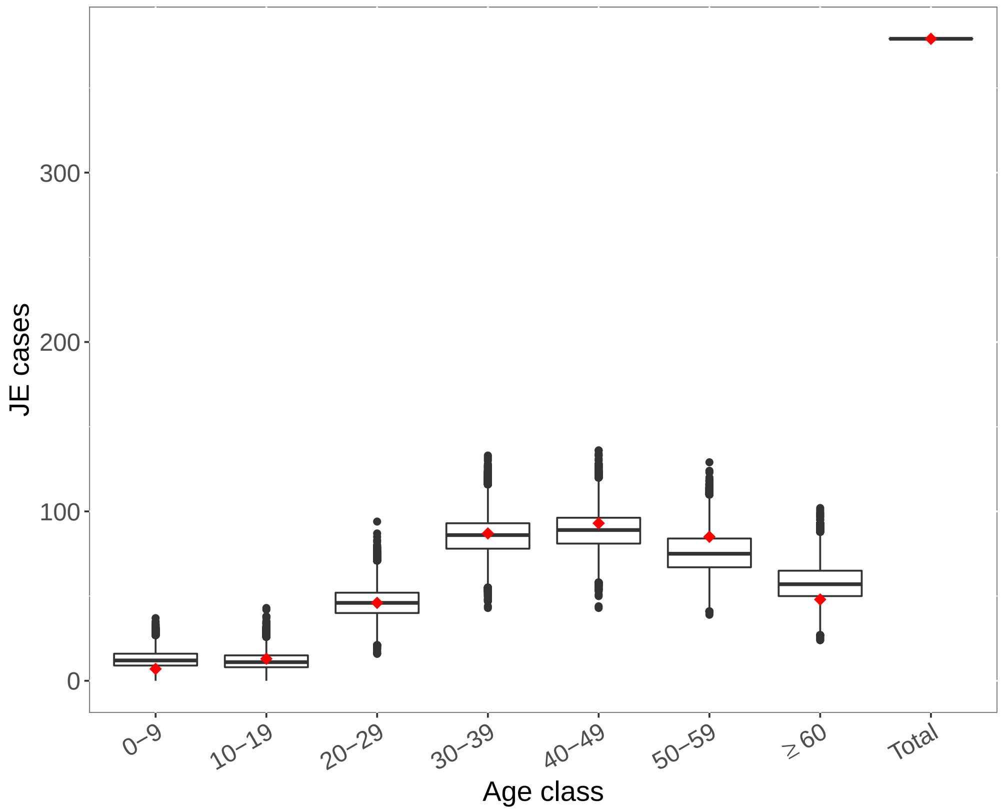


**SI Fig 28.** Observed versus predicted number of JE cases per age class in Taiwan based on dataset from [61]. Boxplots represent predicted number of cases per age class based on draws from the joint posterior distribution of FOI and vaccination coverage (if included) estimates. Red diamonds represent the observed number of cases.

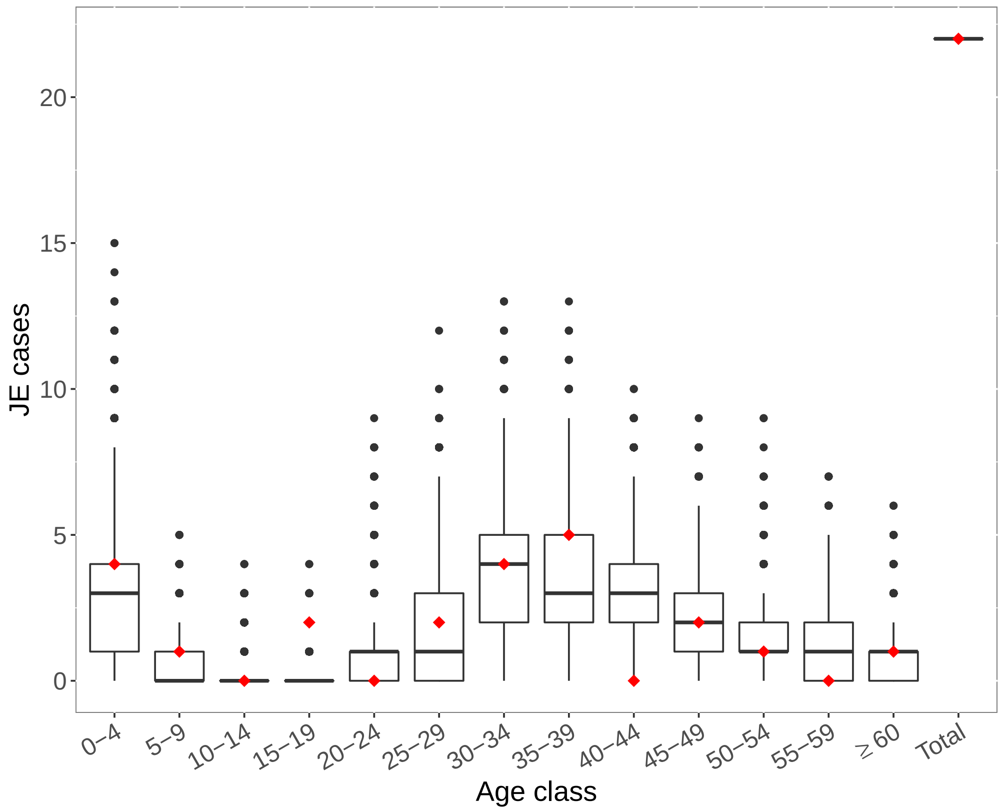


**SI Fig 29.** Observed versus predicted number of JE cases per age class in Thailand based on dataset from [62]. Boxplots represent predicted number of cases per age class based on draws from the joint posterior distribution of FOI and vaccination coverage (if included) estimates. Red diamonds represent the observed number of cases.

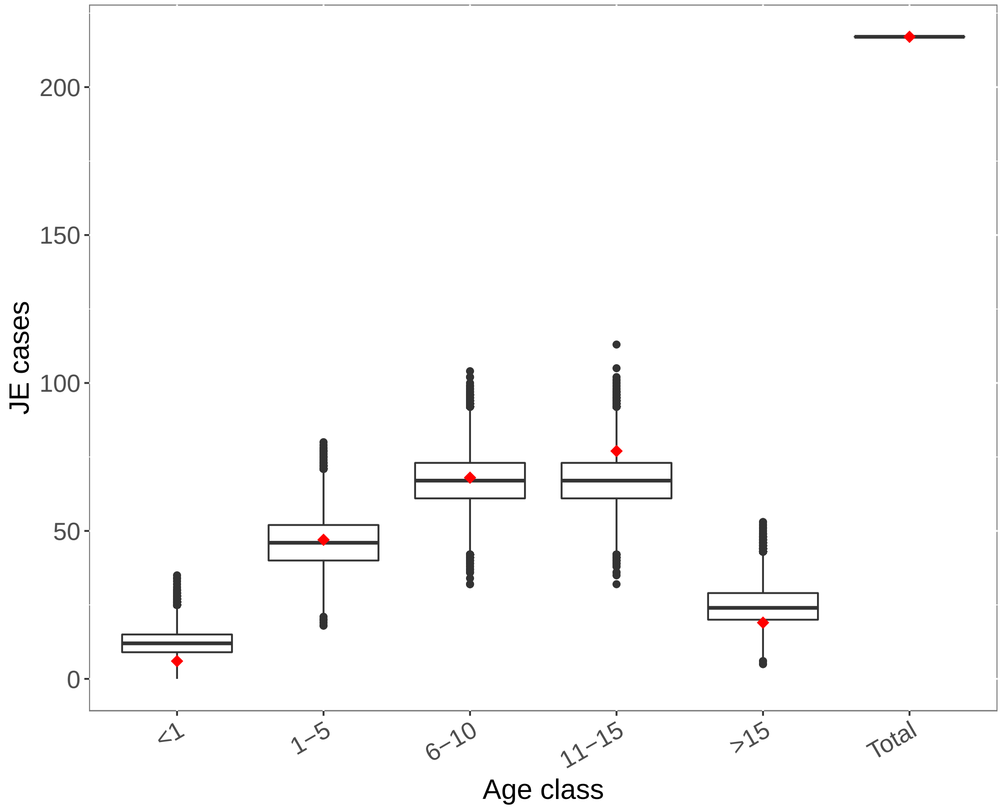


**SI Fig 30.** Observed versus predicted number of JE cases per age class in Vietnam based on dataset from [63]. Boxplots represent predicted number of cases per age class based on draws from the joint posterior distribution of FOI and vaccination coverage (if included) estimates. Red diamonds represent the observed number of cases.

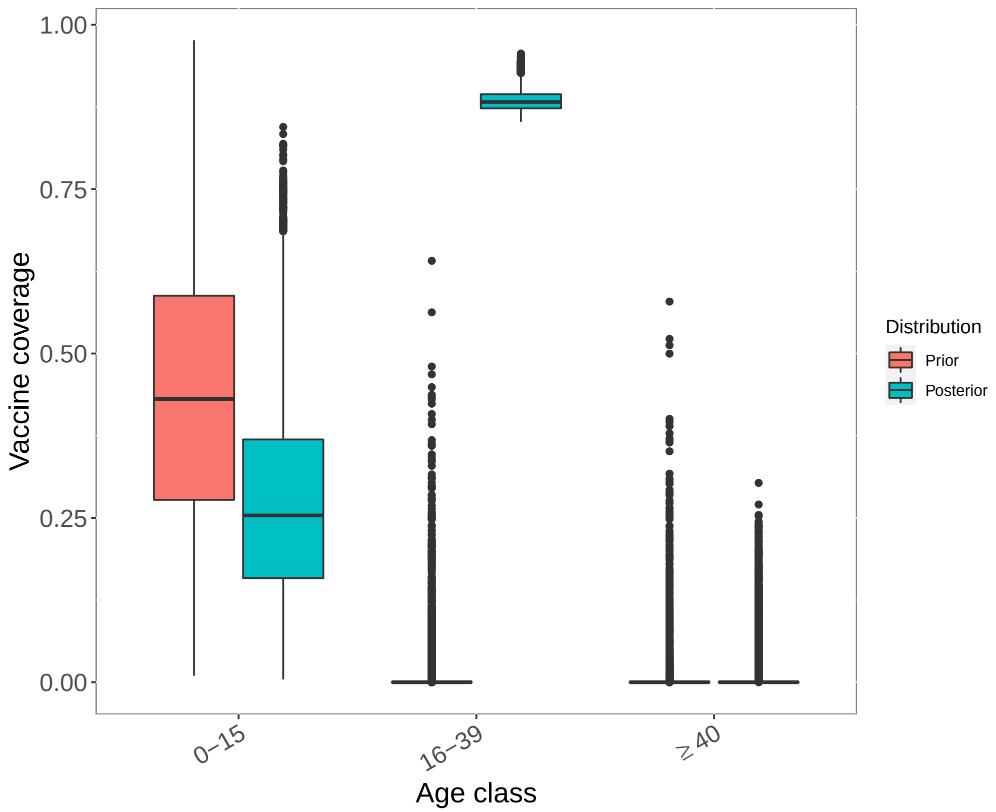


**SI Fig 31.** Estimated vaccination coverage by age group for China based on age-specific incidence data from Li et al. [43]. Vaccination coverage data used to generate prior distributions was taken from reference(s) listed in SI Table 1.

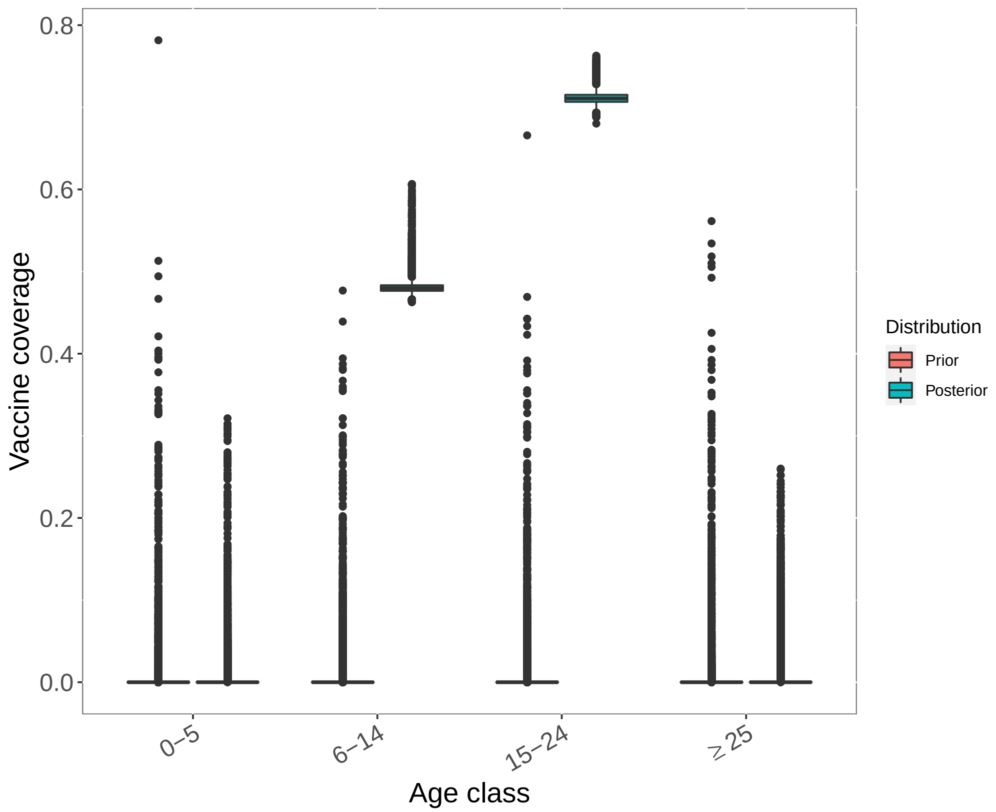


**SI Fig 32.** Estimated vaccination coverage by age group for China based on age-specific incidence data from Wang et al. [44]. Vaccination coverage data used to generate prior distributions was taken from reference(s) listed in SI Table 1.

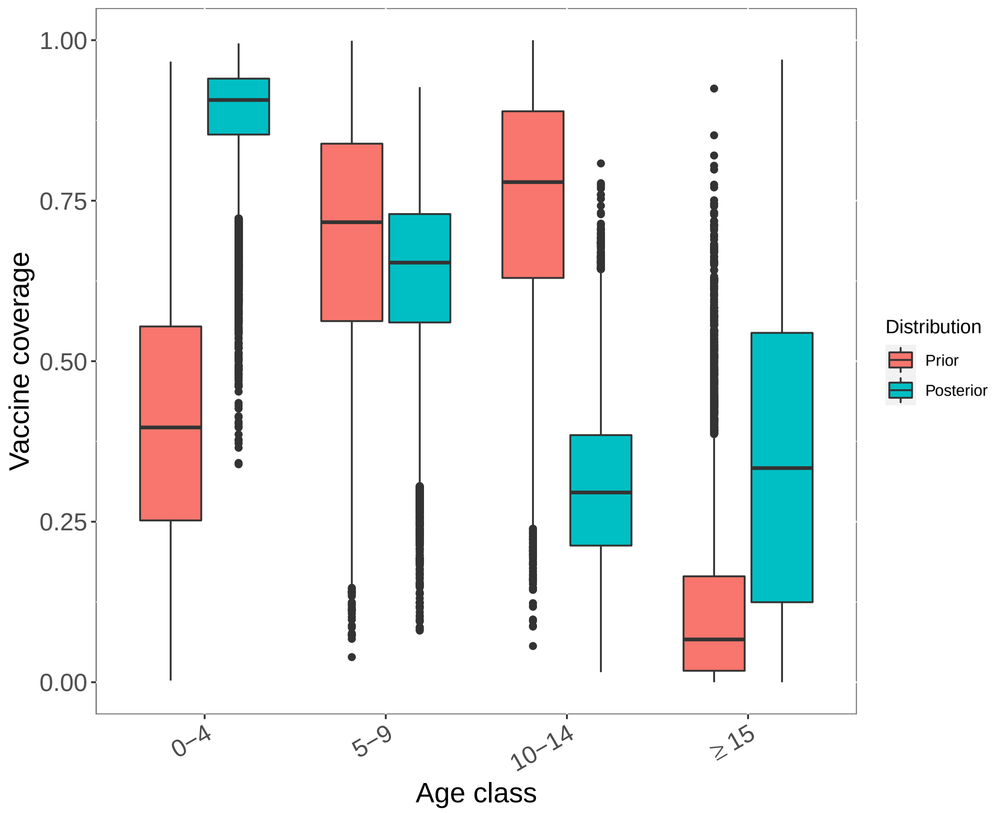


**SI Fig 33.** Estimated vaccination coverage by age group for India based on age-specific incidence data from Ranjan et al. [45]. Vaccination coverage data used to generate prior distributions was taken from reference(s) listed in SI Table 1.

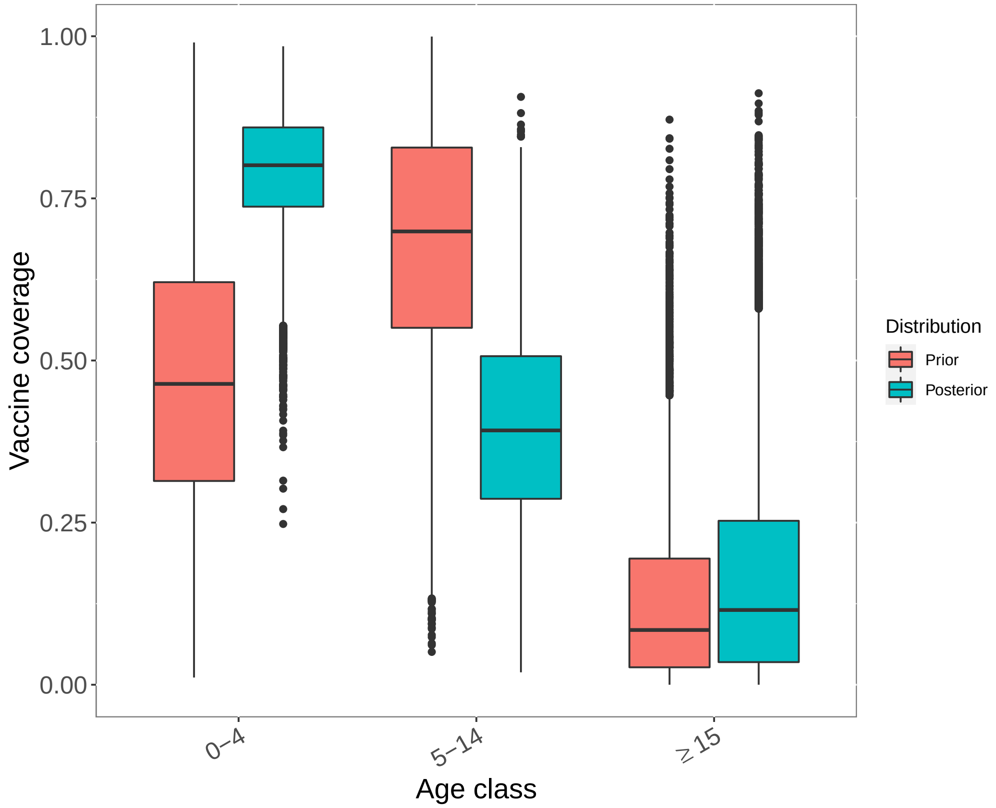


**SI Fig 34.** Estimated vaccination coverage by age group for India based on age-specific incidence data from Jain et al. [47]. Vaccination coverage data used to generate prior distributions was taken from reference(s) listed in SI Table 1.

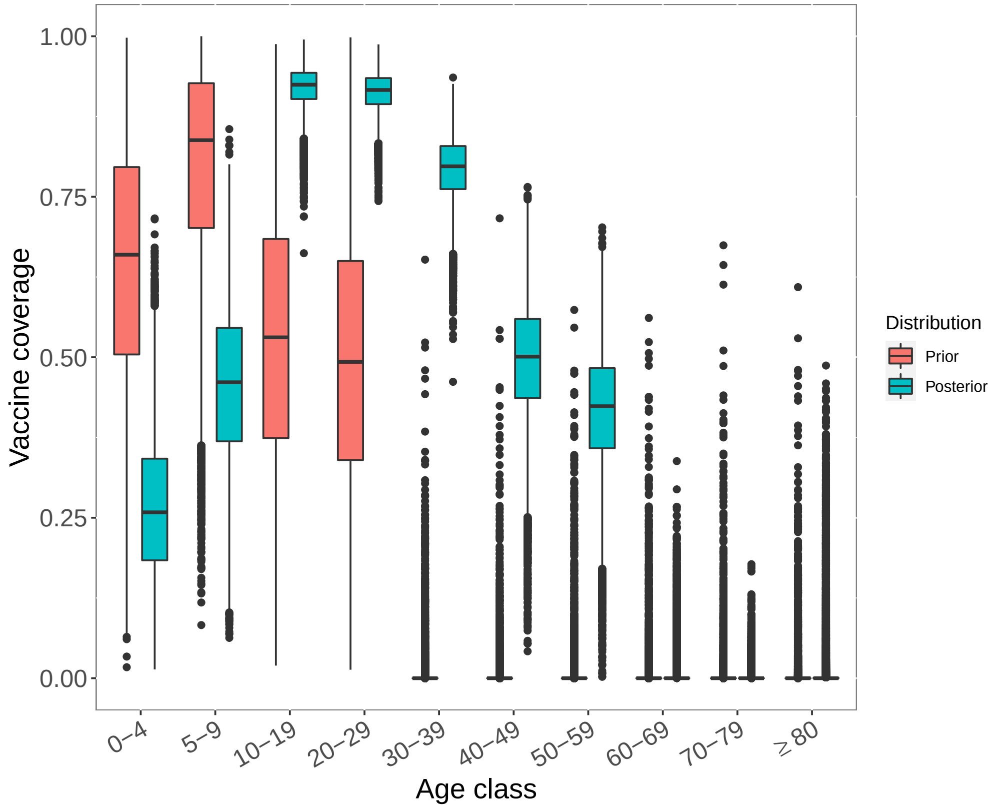


**SI Fig 35.** Estimated vaccination coverage by age group for Japan based on age-specific incidence data from Arai et al. [49]. Vaccination coverage data used to generate prior distributions was taken from reference(s) listed in SI Table 1.

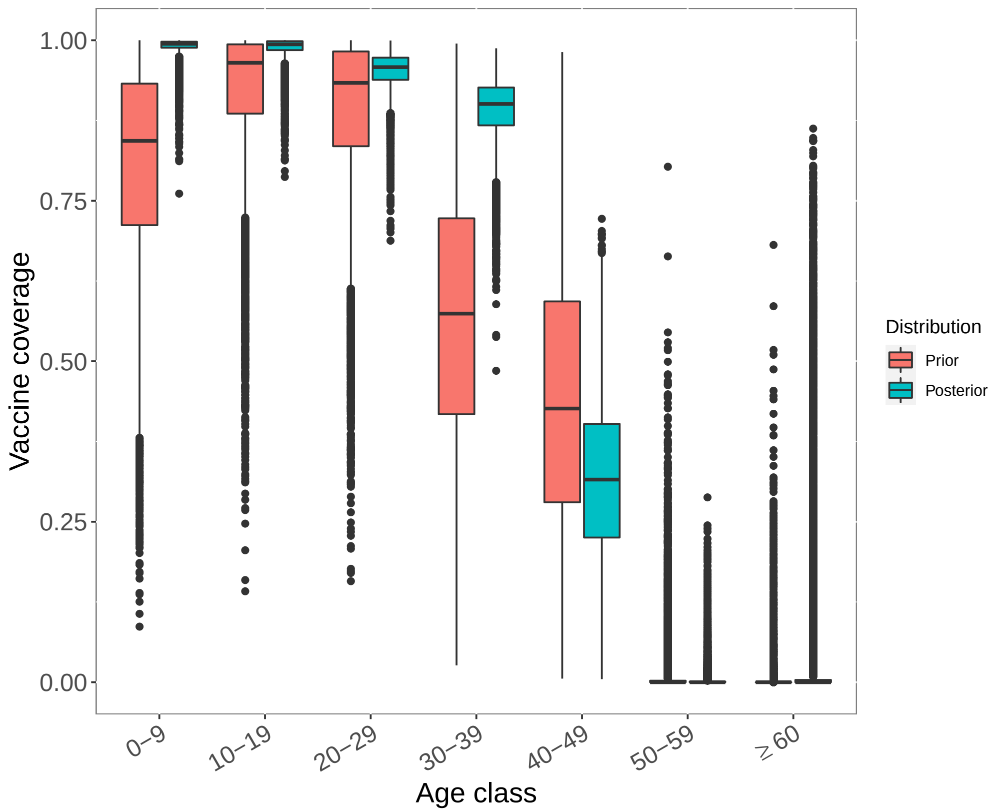


**SI Fig 36.** Estimated vaccination coverage by age group for South Korea based on age-specific incidence data from Lee et al. [59]. Vaccination coverage data used to generate prior distributions was taken from reference(s) listed in SI Table 1.

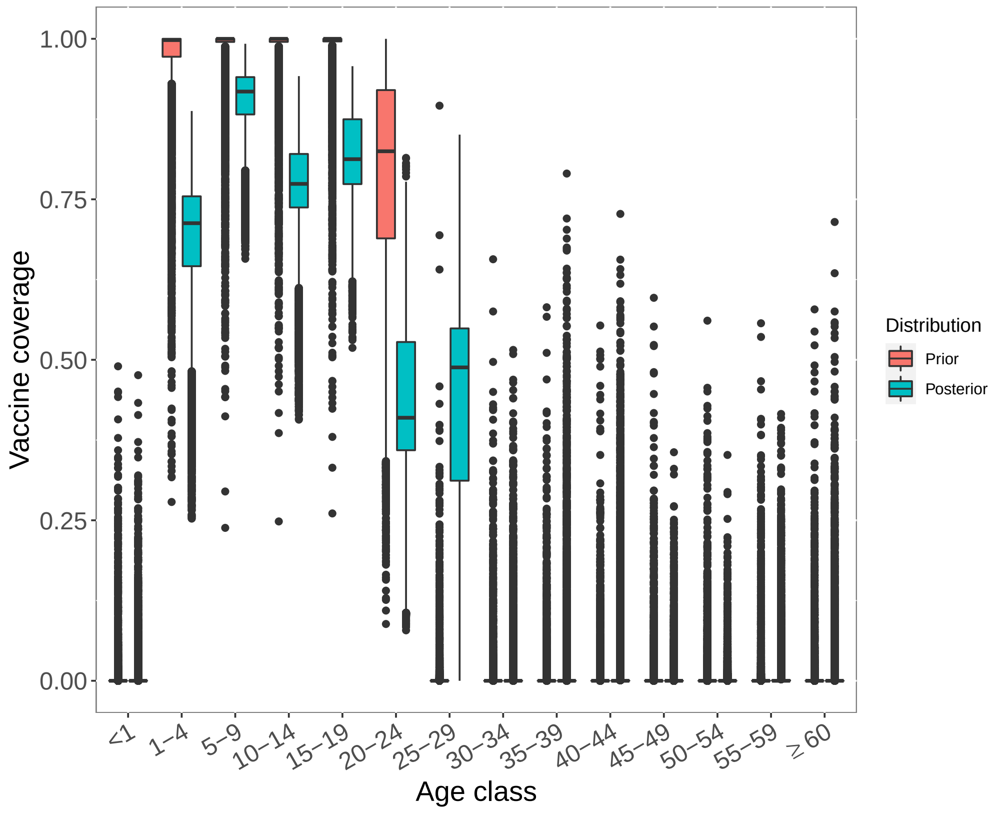


**SI Fig 37.** Estimated vaccination coverage by age group for Sri Lanka based on age-specific incidence data from the Epidemiology Unit of the Sri Lankan Ministry of Health [60]. Vaccination coverage data used to generate prior distributions was taken from reference(s) listed in SI Table 1.

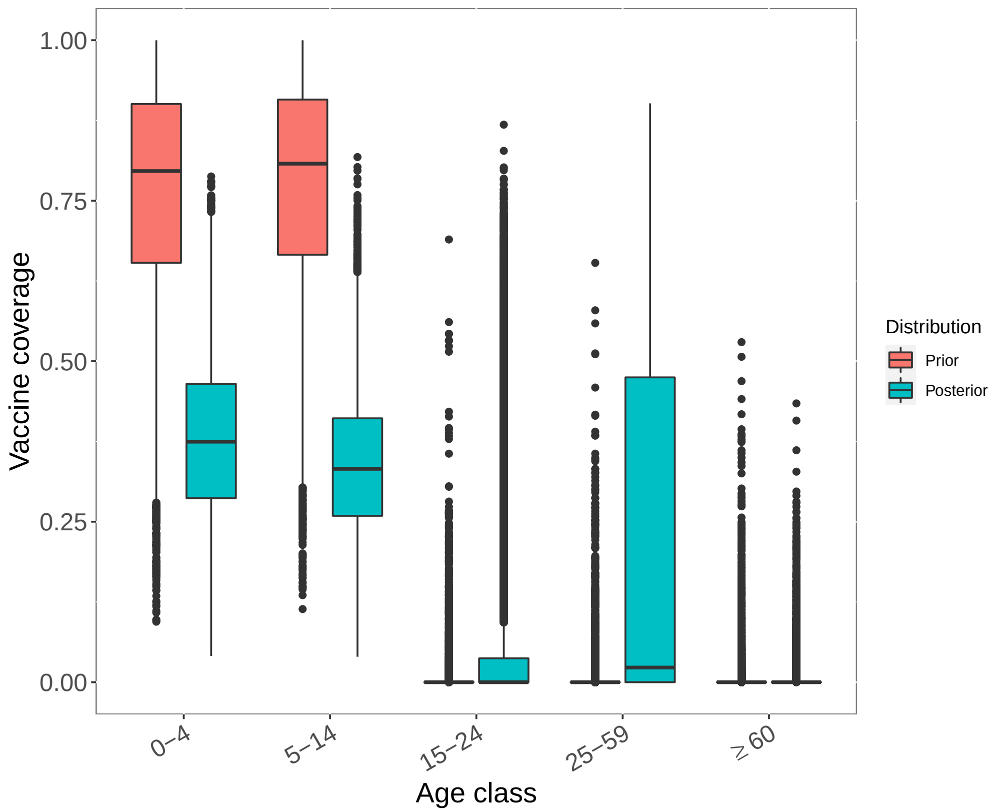


**SI Fig 38.** Estimated vaccination coverage by age group for Malaysia based on age-specific incidence data from Mustapa et al. [51]. Vaccination coverage data used to generate prior distributions was taken from reference(s) listed in SI Table 1.

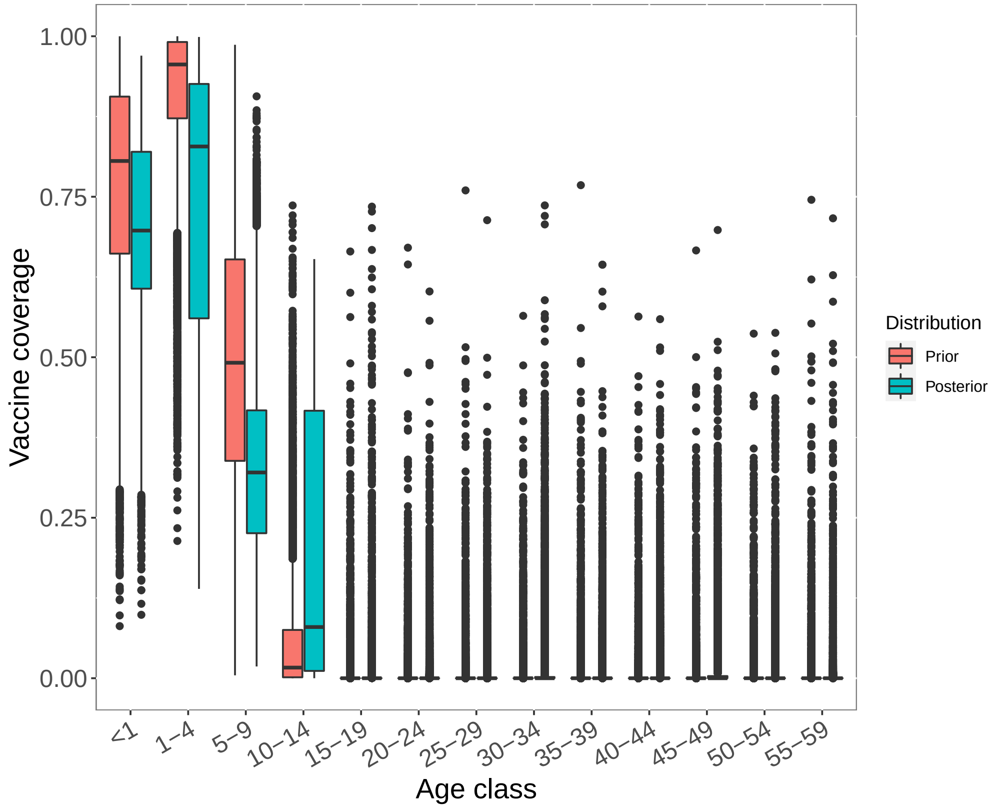


**SI Fig 39.** Estimated vaccination coverage by age group for Thailand based on age-specific incidence data from Olsen et al. [62]. Vaccination coverage data used to generate prior distributions was taken from reference(s) listed in SI Table 1.

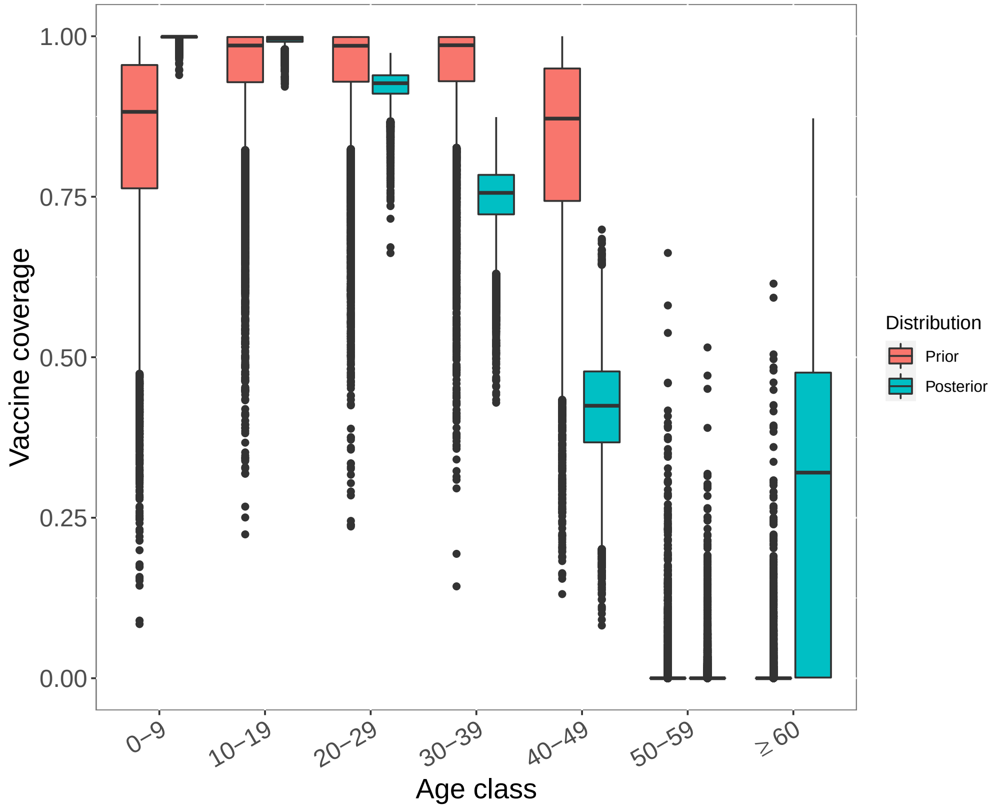


**SI Fig 40.** Estimated vaccination coverage by age group for Taiwan based on age-specific incidence data from Chang et al. [61]. Vaccination coverage data used to generate prior distributions was taken from reference(s) listed in Table 1.

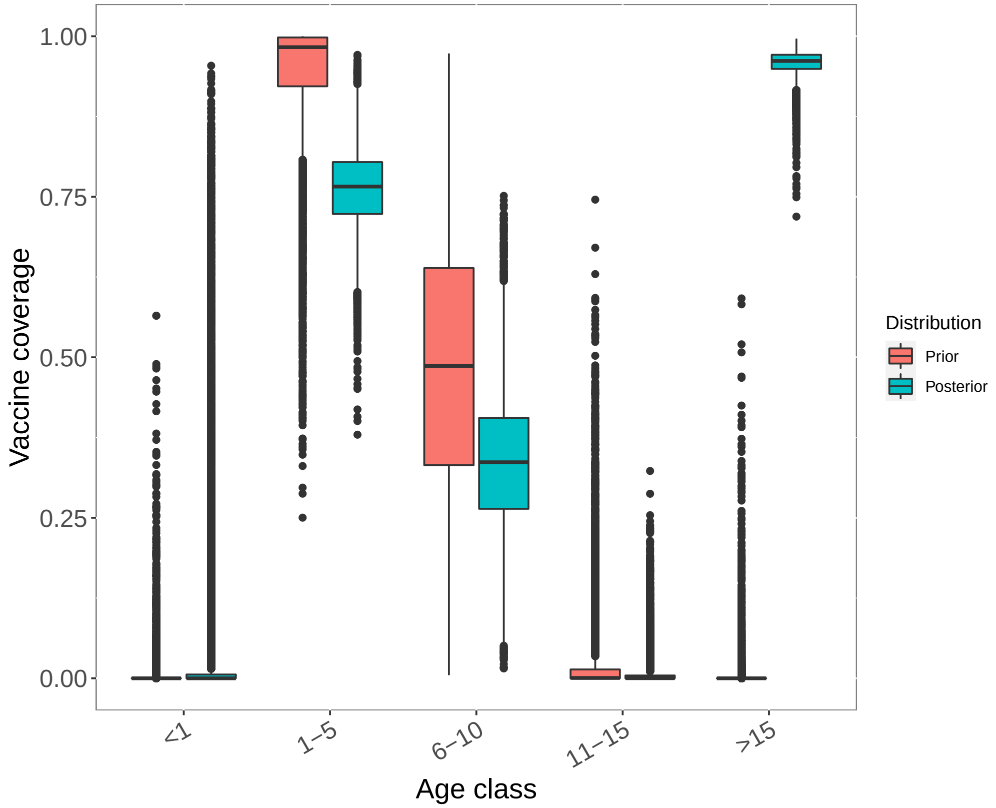


**SI Fig 41.** Estimated vaccination coverage by age group for Vietnam based on age-specific incidence data from Yen et al. [63]. Vaccination coverage data used to generate prior distributions was taken from reference(s) listed in Table 1.
